# Scalable cell-state-resolved TWAS identified dynamic autoimmune disease genes along functional programs

**DOI:** 10.64898/2026.11.20.26368605

**Authors:** Suyang Yu, Jiayu Chen, Saori Sakaue, Wei Sun, Eardi Lila, Guanghao Qi

## Abstract

Transcriptome-wide association studies (TWAS) are widely used to prioritize genes for complex diseases. TWAS typically trains models on expression quantitative disease locus (eQTL) reference data to predict gene expression from genotype, then tests for association between the predicted expression and disease. With the rapid growth of single-cell eQTL data, there is a pressing need for methods that can fully leverage cell-level information. We previously developed TWiST, the first method for TWAS at cell-state resolution. TWiST is powerful, but training its prediction model is computationally expensive. Here we propose Fast-TWiST, an accelerated and scalable version of TWiST that can analyze hundreds of thousands to millions of cells. Fast-TWiST computes donor-level summary statistics that retain all the information needed to fit the cell-level model, so numerical optimization runs over donors rather than individual cells. In simulations and real-data evaluations, Fast-TWiST was more than 10 times faster than TWiST while matching its statistical performance. Applying the method to the OneK1K data, we identified 3,092 dynamic gene-disease associations that depend on biologically interpretable gene programs, including cytotoxicity, immediate early gene (IEG), BCL2/FAM13A, T helper 1-like (Th1-Like), and resting T helper 2 (Th2-Resting). Independent eQTL datasets and single-cell multiome data supported cell-state-specific regulatory effects that may be driving these findings.

## Introduction

Transcriptome-wide association studies (TWAS) prioritize disease-relevant genes by integrating genetic associations with expression quantitative trait loci (eQTLs). These methods are motivated by the hypothesis that genetic variants influence disease risk through their regulatory effects on gene expression. Since their introduction,^1,2^ TWAS have been applied to a wide range of traits and diseases,^3–7^ identifying thousands of gene-disease associations. The statistical workflow of TWAS involves two stages: first, training gene expression prediction models in an eQTL reference panel; and second, integrating these models with genome-wide association study (GWAS) to test gene-level associations with traits. Conventional TWAS typically use bulk eQTL reference panels, such as the Genotype-Tissue Expression (GTEx) project,^8^ the eQTLGen Consortium,^9^ and the Depression Genes and Networks (DGN).^10^ Although these reference panels can provide tissue-specific information, bulk tissues are comprised of a mixture of cell types, which can obscure cell-type-specific regulatory effects.

Single-cell eQTL studies have expanded rapidly in recent years,^11–18^ enabling the characterization of genetic regulatory effects at cellular resolution. Several methods, including scPrediXcan,^19^ scTWAS,^20^ MOD-scTWAS,^21^ and Abe et al,^22^ have been developed to perform TWAS using single-cell eQTL data at the cell-type level. However, substantial heterogeneity can exist within a cell type, including continuous variation of cell states that may shape genetic regulatory effects.^11–13,15,23^ We previously developed TWiST,^24^ the first method for performing TWAS at cell-state resolution. Application of TWiST identified hundreds of genes with disease associations that varied along the naïve-to-memory trajectories of T and B cells.^24^

A major limitation of TWiST is its computational cost. Its Stage-1 prediction models are fitted using single cells or meta-cells, which ranges from tens to hundreds of thousands.^24^ These models use B-splines to model SNP effects on gene expression as functions of cell states, hence expanding the design matrix several times compared to models where SNP effects are constant. In the original study, we could not fit models directly to individual cells due to computational constraints. Instead, we aggregated approximately every five cells into a meta-cell to reduce the computational burden. This computational challenge is shared by another cell-state-level TWAS method, pt-TWAS,^25^ which also adopted the meta-cell approach. As single-cell eQTL studies continue to grow, even this approach is likely to encounter substantial computational bottlenecks.

Here, we introduce Fast-TWiST, an accelerated framework for cell-state-resolved TWAS. Its primary approach is a compressed functional lasso (compressed FLASSO) method that aggregates cell-level information into donor-level summary statistics and performs optimization using these compressed quantities. Because the resulting computations scale with the number of donors rather than the number of cells, this approach can accommodate increasingly large single-cell eQTL datasets. We also introduce an alternative approach based on functional principal component analysis (FPCA), which uses a set of data-driven basis functions to model the curves of gene expression and genetic regulatory effects. To combine the strengths of these approaches, we aggregate their association evidence using the Cauchy combination test (CCT)^26^.

Through simulation studies and real data analysis, we demonstrated that Fast-TWiST was more than ten times faster than the original TWiST, with modest gains in statistical power through the CCT. We first revisited the cell-state-resolved TWAS analyses of three cell types from the original TWiST study, achieving shorter runtimes and robust replication rates. We then applied Fast-TWiST to cell states defined by five biologically interpretable gene expression programs (GEPs)^27^, identifying more than 1,000 genes with cell-state-dependent associations with autoimmune diseases. We further characterized the patterns of these associations and conducted validation analyses using independent datasets.

## Results

### Overview of Fast-TWiST

TWiST^24^ conducts TWAS at cell-state resolution. It models gene expression and its effect on disease as functions of a continuous cell state *t* (Figure 1a). In Stage 1, the genetically regulated expression (GReX) of donor *i, v*_*i*_(*t*), enters the log-mean in a Poisson model for UMI counts. The GReX is a linear function of genotypes whose coefficients vary with *t*:

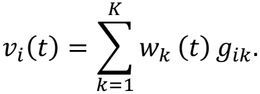

**Figure 1.**
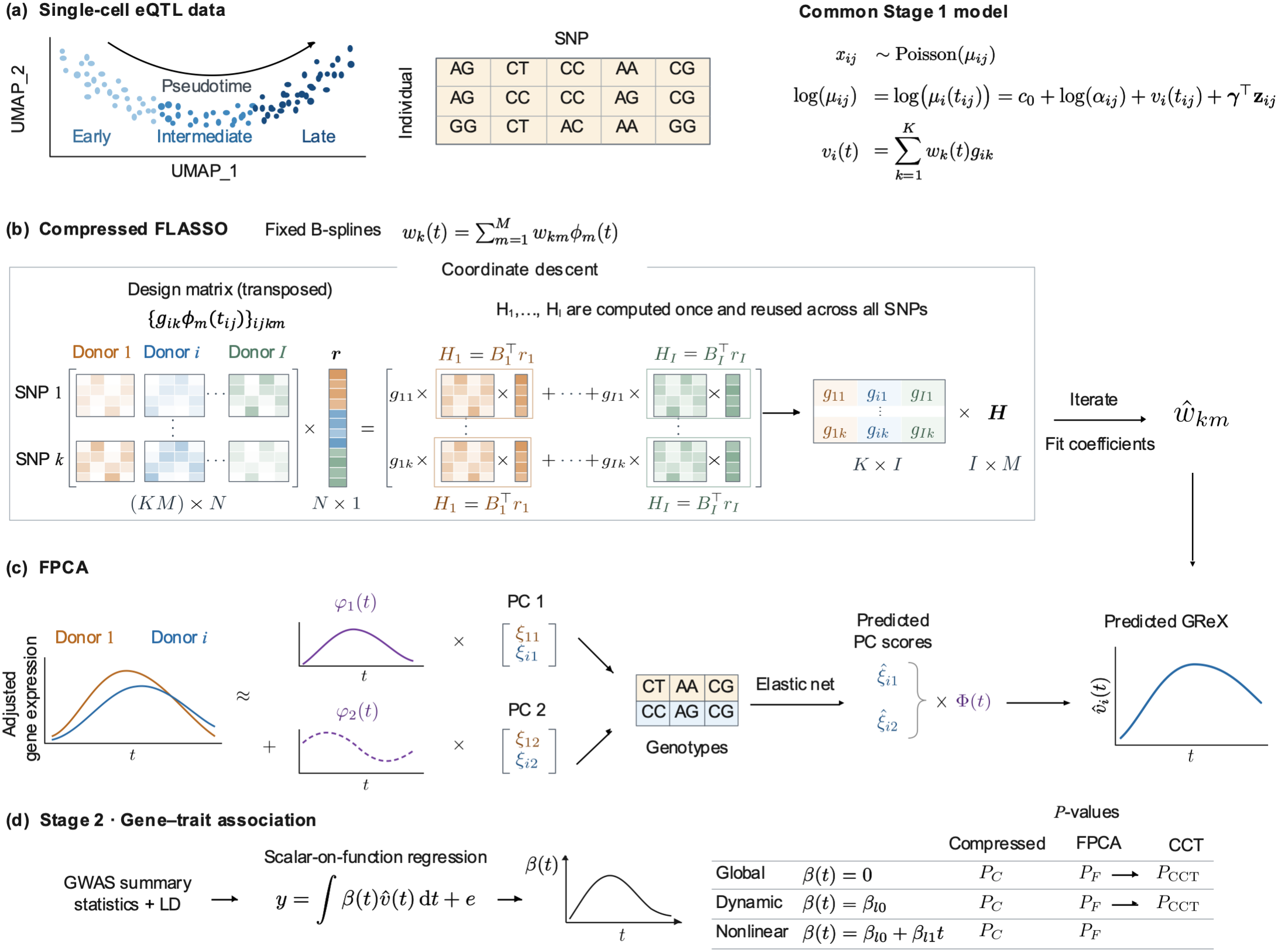
Overview of Fast-TWiST. (a) Single-cell eQTL data consist of single-cell RNA-seq data for gene expression and SNP genotypes of the same individuals. Cells can be ordered from early to late along a cell state variable *t* (e.g., pseudotime; each dot in the UMAP is a cell). In the Stage-1 model shared by both methods, the UMI count *x*_*ij*_ of cell *j* from individual *i* is modeled with a Poisson distribution whose mean µ_*ij*_ depends on an intercept *c*_0_, library size α_*ij*_, covariates **z**_*ij*_ with effects γ, and the genetically regulated expression (GReX) *v*_*i*_(*t*), a continuous curve over pseudotime *t*. GReX is a linear function of the genotypes *g*_*ik*_ of *K* cis-SNPs weighted by SNP effects *w*_*k*_(*t*). (b) Compressed FLASSO represents each *w*_*k*_(*t*) with *M* fixed B-spline functions *ϕ*_1_(*t*) and estimates the coefficients *w*_*km*_ by coordinate descent. Each update multiplies the design matrix by the residuals **r** of all *N* cells. Because cells from the same individual (Indiv) share the same genotype, this product can be computed from individual-level summaries 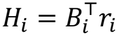, where *B*_*i*_ and *r*_*i*_ are the B-spline values and residuals for the cells of individual *i*. The summaries, stacked in **H**, are shared by all SNPs, so the computation scales with the number of individuals *I* instead of *N*. (c) FPCA approximates the adjusted gene expression curve of each individual with a few data-driven functions *φ*_1_(*t*), *φ*_2_(*t*), …, and individual-specific scores ξ_*i*1_, ξ_*i*2_, …. The scores are predicted from genotypes using elastic net and then combined with Φ(*t*) to give the predicted GReX 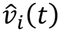. (d) The predicted GReX from either method is linked to a trait *y* through a scalar-on-function regression model with residual *e*, using GWAS summary statistics and linkage disequilibrium (LD). The estimated effect *β*(*t*) is a continuous curve and is used for global, dynamic, and nonlinear tests. The P values from compressed FLASSO (*P*_*C*_) and FPCA (*P*_*F*_) are combined into *P*_CCT_ using the Cauchy combination test (CCT).

TWiST then models each coefficient function *w*_*k*_(*t*) as a B-spline function, *w*_*k*_(*t*) = ∑_*m*_ *w*_*km*_ϕ_*m*_(*t*). This turns the problem to a standard Poisson regression with an *N* × *KM* design matrix, where *N, K*, and *M* are the number of cells, cis-SNPs, and B-spline bases. This large design matrix is used repeatedly during numerical optimization, incurring large computational cost.

Our new method, Fast-TWiST, speeds up Stage-1 model training with two algorithms (Figure 1). The primary algorithm, compressed FLASSO, fits the same model as TWiST with a faster coordinate descent algorithm (Figure 1b). The main cost of each iteration is the cross-product *G* ^⊤^ ***r*** of the design matrix (*G*) and working residual (***r***), which is proportional to ***x*** − µ at the current iteration (Figure 1b). This product sums across all cells and covers all SNPs, and is computed every iteration and every cross-validation fold. It therefore dominates the computational time when there are many cells. The compressed FLASSO exploits the fact that genotypes are constant across all cells from the same donor, so for donor *i* and SNP *k* the donor’s contribution is 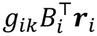. Here *B*_*i*_ is the *J*_*i*_ × *M* matrix of B-spline bases evaluated at the donor’s *J*_*i*_ cells and ***r***_*i*_ is the matching residual vector. The product 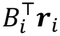 does not depend on SNP *k*, so it is computed from the cells once per iteration and shared by all the SNPs. This turns the cell-level work per iteration from *O*(*NKM*) into *O*(*NM*), plus an inexpensive donor-level step.

A second algorithm uses FPCA (Figure 1c). It represents each donor’s expression trajectory as a linear combination of data-driven basis functions, weighted by donor-specific FPC scores. These scores are regressed on genotypes to build the prediction model. Because the optimization runs on donor-level summaries instead of cell-level counts, it is much faster than count-based fitting. The predicted scores are then combined with the FPC basis functions to give the predicted GReX.

Both methods produce GReX predictions for the Stage-2 association analysis. Finally, we combine the P-values from the two algorithms with the Cauchy combination test (CCT) to leverage their complementary strengths (Figure 1d). See Methods and Supplementary Methods S1 for details.

### Simulation studies

We evaluated the computational efficiency and statistical performance of Fast-TWiST using the simulation design developed for TWiST (see Methods). Single-cell expression data were simulated using a Poisson regression model under two gene-expression noise levels: 10x noise and no noise. GWAS summary statistics were generated under four types of gene-trait effect curves, *β*(*t*): null, constant, unimodal, and reverse (Figure 2). We compared Fast-TWiST with TWiST and pseudobulk TWAS models trained using elastic net or lasso. Fast-TWiST reduced Stage 1 training time by more than 10-fold across all simulation settings (Figure 2a and

**Figure 2.**
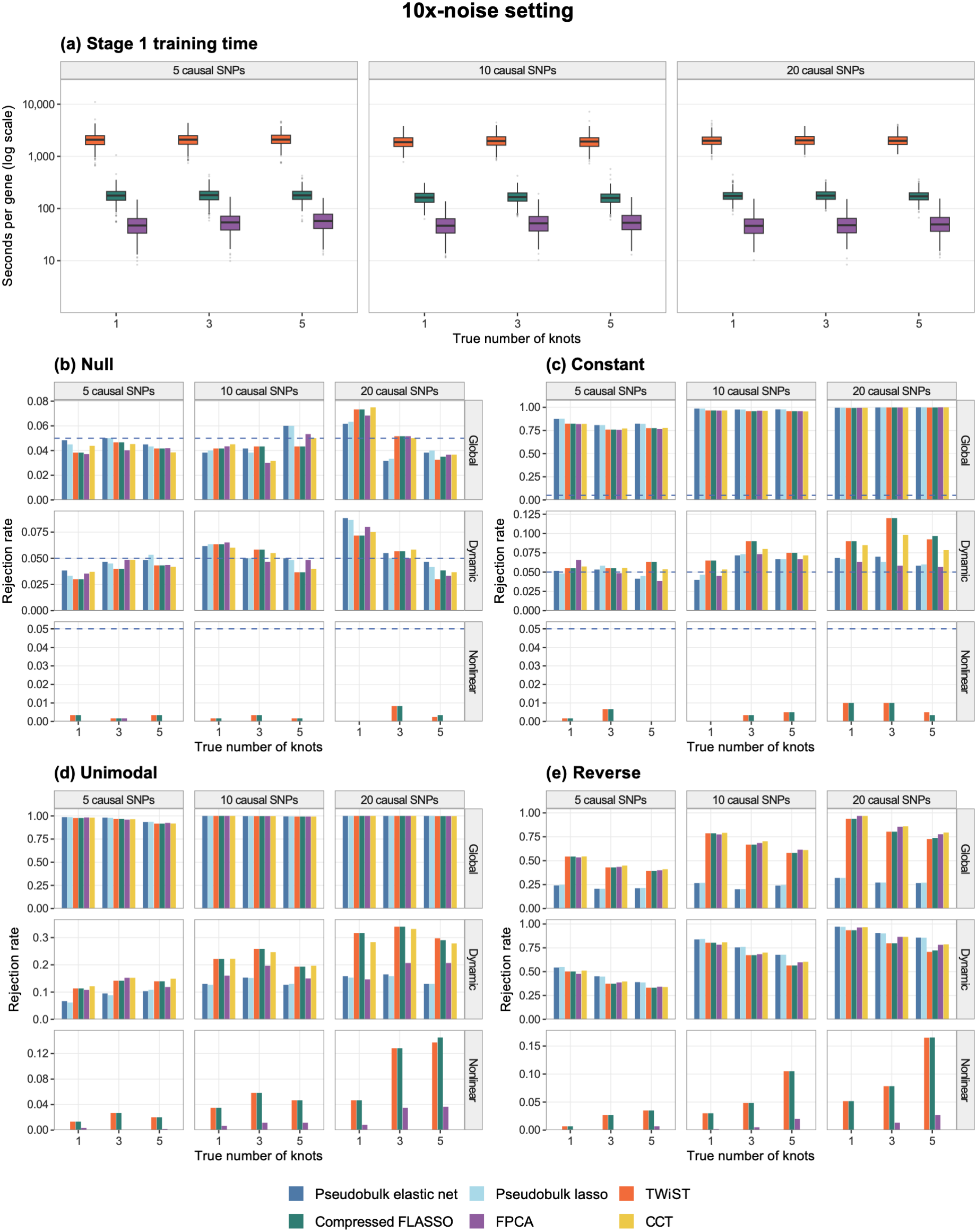
Computation time, type I error, and power observed in simulation studies. (a) Stage-1 training time per gene for TWiST, compressed FLASSO, and FPCA. (b) Type I error for global, dynamic, and nonlinear tests in null simulations. (c) Power of the global test and type I error of the dynamic and nonlinear tests in the constant scenario. (d-e) Power in the unimodal and reverse scenarios. See Methods for description of simulation setting. Six methods were compared: pseudobulk elastic net, pseudobulk lasso, TWiST, compressed FLASSO, FPCA, and the Cauchy combination test (CCT) of compressed FLASSO and FPCA. The number of causal SNPs in the cis-region of each gene was 5,10,20. True SNP effects on expression and the true gene effect on the trait, *β*(*t*) are B-spline functions between 0 and 1 with 1, 3, or 5 knots. Independent Gaussian noise was added to the log mean of expression, with variance approximately ten times the variance of the genetic component. Blue dashed lines represent significant threshold p = 0.05.

Supplementary Figure S1). In the 10x-noise setting (see Methods for description), median training time across all simulation settings was 2,004 seconds per gene for TWiST, 172 seconds for compressed FLASSO, and 50 seconds for FPCA (Figure 2a). Without added noise (Supplementary Figure S1), the corresponding median time was 1,653 seconds for TWiST, 141 seconds for compressed FLASSO, and 48 seconds for FPCA. The training time was approximately identical across the numbers of causal SNPs and B-spline knots.

With improved computational efficiency, Fast-TWiST had similar type I error and power to TWiST across simulation settings, both of which outperformed pseudobulk methods in most setting (Figure 2b-e). Under the null scenario, type I error rates for the global and dynamic tests were close to the nominal level of 0.05, whereas the nonlinear tests were conservative (Figure 2b). Under constant effects, all methods had high power for the global test (Figure 2c). However, the dynamic tests showed slightly elevated type I error in some settings, particularly for TWiST and compressed FLASSO with 20 causal SNPs. FPCA controlled type I error for dynamic tests across most scenarios, and the CCT had less type I error than TWiST and compressed FLASSO but slightly more than FPCA.

When the effect varied over pseudotime (Figure 2d and 2e, unimodal and reverse), Fast-TWiST outperformed the pseudobulk methods. In the unimodal scenario (Figure 2d), all methods had high power for the global test. For the dynamic test, TWiST and compressed FLASSO had similar power and both were more powerful than the pseudobulk methods. FPCA was more powerful than the pseudobulk methods but considerably less powerful than TWiST and compressed FLASSO. The CCT had similar power to TWiST overall. For the nonlinear test, compressed FLASSO had similar power to TWiST, while FPCA had no power. We did not apply the CCT to the nonlinear test because its null distribution is irregular (see Methods and Supplementary Methods), and FPCA had no power. In the reverse scenario (Figure 2e), the main advantage of the functional methods was instead observed in the global test. TWiST, compressed FLASSO, and FPCA were all several times more powerful than the pseudobulk methods. All methods had similar power for dynamic test. The CCT had slightly higher power than TWiST and compressed FLASSO for both the global and dynamic tests. In the no-noise setting, CCT improved power over compressed FLASSO for the dynamic test in the unimodal scenario, whereas no gains were observed in the reverse scenario (Supplementary Figure S1).

### Performance of Fast-TWiST in the OneK1K data

To evaluate the performance of Fast-TWiST in real data, we applied the method to the three immune cell types from the OneK1K study^12^: CD4^+^ T cells, CD8^+^ T cells, and B cells. We used pseudotime to represent the transition from naïve to memory cells. Due to the large number of cells (547,509 CD4^+^ T cells, 68,889 CD8^+^ T cells, and 125,746 B cells from 980 individuals), it was computationally infeasible to train the original TWiST GReX prediction models on single cells. Therefore, we trained TWiST Stage-1 models on meta-cells (pseudobulk sample of approximately 5 cells),^24^ which had median runtime of 69.82, 27.58, and 30.60 minutes per gene for CD4^+^ T cells, CD8^+^ T cells, and B cells, respectively. In contrast, Fast-TWiST successfully trained prediction models directly on single cells. The median runtime per gene was 2.33, 5.96, and 3.64 minutes for compressed FLASSO and 1.04, 0.66, and 0.76 minutes for FPCA in CD4^+^ T cells, CD8^+^ T cells, and B cells, respectively (Figure 3a). We then applied the models to re-analyze the seven autoimmune diseases analyzed in the TWiST paper: rheumatoid arthritis (RA),^28^ systemic lupus erythematosus (SLE),^29^ Crohn’s disease (CD),^30^ inflammatory bowel disease (IBD),^30^ multiple sclerosis (MS),^31^ type 1 diabetes (T1DM),^32^ and ankylosing spondylitis (AS).^32^ For CD4^+^ T cells and B cells, we observed stronger signal enrichment for CCT and compressed FLASSO for both the global and dynamic tests, with their QQ plots dominating that of TWiST among the most significant genes (Figure 3b and 3c). For CD8^+^ T cells, the QQ plots of CCT and compressed FLASSO were higher than that of TWiST for genes of intermediate significance, but lower for the most significant genes.

**Figure 3.**
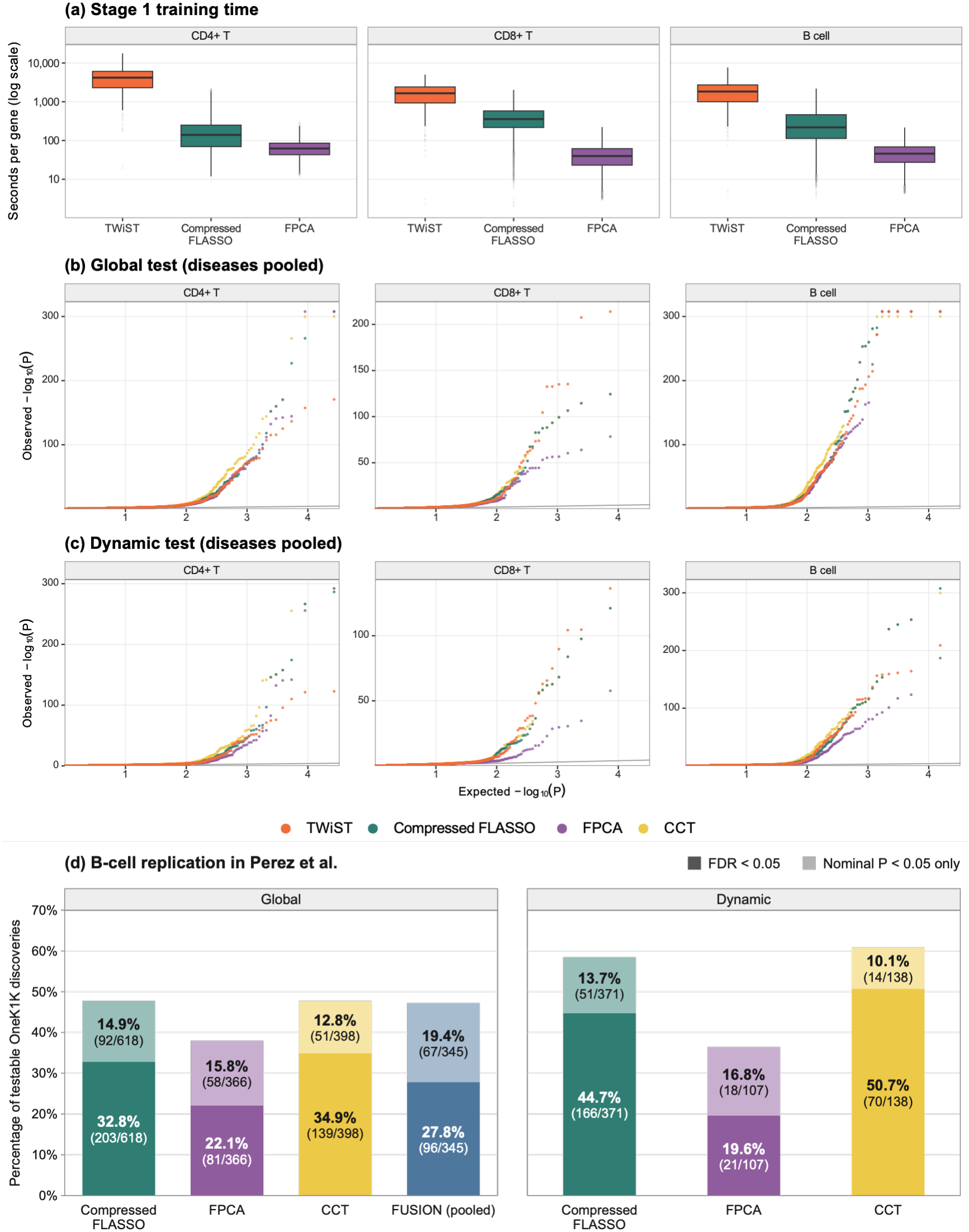
Evaluation of Fast-TWiST in the OneK1K data. (a) Stage-1 training time per gene for TWiST on meta-cells and Fast-TWiST (compressed FLASSO and FPCA) on single-cells. Each meta-cell is an aggregate of 5 single cells on average. (b-c) QQ plots for the (b) global and (c) dynamic tests across seven autoimmune disease and three immune cell types. For each combination of cell type, disease and test, the common set of gene with valid p-values for all methods are included in the QQ plot. Diseases are pooled, each cell type is plotted separately. (d) Percentage of Fast-TWiST findings for OneK1K B cells replicated in independent datasets. For each gene-disease association identified in the OneK1K analysis (FDR<0.05 for each disease-method-cell type combination), we checked whether it was significant in the Fast-TWiST analysis of Perez et al and independent GWAS summary statistics. Replication rates were computed with the number of genes that were significant in OneK1K and received a Stage-2 P-value in Perez et al as the denominator. FUSION was applied to the pseudobulk samples of four B cell subtypes (naïve, memory, pseudotime≥0.5, pseudotime<0.5). Replicated analyses were conducted for each subtype. All subtypes were combined to compute the replication rate.

Next, we evaluated whether these findings can be replicated by applying Fast-TWiST to independent datasets: Perez et al single-cell eQTL dataset^14^ and independent GWAS summary statistics for the diseases (see Methods for details). Due to differences in cell type annotation between the OneK1K and Perez et al, we focused on B cells for which the naïve-memory cells are annotated consistently (see Methods for more details, Supplementary Figure S2). After training the Stage-1 model on Perez et al, we obtained 7042 testable genes for compressed FLASSO, 6805 testable genes for FPCA, defined as those with at least 3 SNPs with non-zero coefficients in the prediction model. The CCT requires the gene to be testable for both FLASSO and FPCA, yielding 4427 testable genes. The intersection between OneK1K significant genes and Perez et al testable genes were used as the denominator to compute replication rates.

Across seven autoimmune diseases, compressed FLASSO replicated 203 of 618 global gene-disease associations (32.8%) and 166 of 371 dynamic associations (44.7%) at FDR < 0.05 in the validation data, and 295 of 618 (47.7%) and 217 of 371 (58.5%) at nominal P<0.05, respectively. FPCA replicated 22.1% for global associations and 19.6% for dynamic associations at FDR<0.05. CCT replicated 139 of 398 global associations (34.9%) and 70 of 138 dynamic associations (50.7%) at FDR < 0.05, and 190 of 398 (47.7%) and 84 of 138 (60.9%) at nominal P<0.05, respectively (Figure 3d). In addition, conventional FUSION models^2^ trained on B-naive, B-memory, early (pseudotime< 0.5) and late (pseudotime≥ 0.5) expression profiles were applied as a further benchmark. Fast-TWiST showed higher replication rates than conventional FUSION at both significance thresholds (Figure 3d): FUSION replicated 27.8% (96/345) of testable discoveries at FDR < 0.05 and 47.2% (163/345) at nominal P < 0.05.

### Fast-TWiST identified disease associations across multidimensional T-cell states

T cells include many subpopulations distinguished by various molecular functions, such as CD4 versus CD8, naïve versus memory, T cell receptor complexes, and specific gene programs.^33^ This complexity cannot be captured by a one-dimensional pseudotime. Therefore, we applied starCAT^27^ to define fine-grained, interpretable cellstates by the usage values of 52 gene expression programs (GEPs) in OneK1K T cells (Supplementary Figure S3). We selected five GEPs with relatively high usage and distinct distributions across the T-cell transcriptional landscape: Cytotoxic, immediate early gene (IEG), BCL2/FAM13A, T helper 1-like (Th1-Like), and resting T helper 2 (Th2-Resting), while excluding components primarily associated with cell-type identity or technical variation (see Methods for more details).

Projection of the five GEPs onto a common UMAP revealed distinct and partially overlapping patterns of GEP usage (Figure 4a). For example, Cytotoxic usage was concentrated in cytotoxic and effector-associated T-cell populations, whereas IEG exhibited a broader distribution across the transcriptional landscape (Figure 4a and Supplementary Figures S4, S5). To characterize their correspondence with conventional cell-type annotations, we ranked T-cell subtypes by their mean GEP usage (Figure 4b). Cytotoxic usage was highest in CD4 cytotoxic and CD8 effector memory cells, whereas IEG was most prominent in CD4 effector memory, CD4 central memory and regulatory T cells. BCL2/FAM13A showed higher usage in CD4 naive and central memory cells, while Th1-Like and Th2-Resting were most prominent in CD4 effector memory and CD8 central memory populations, respectively.

**Figure 4.**
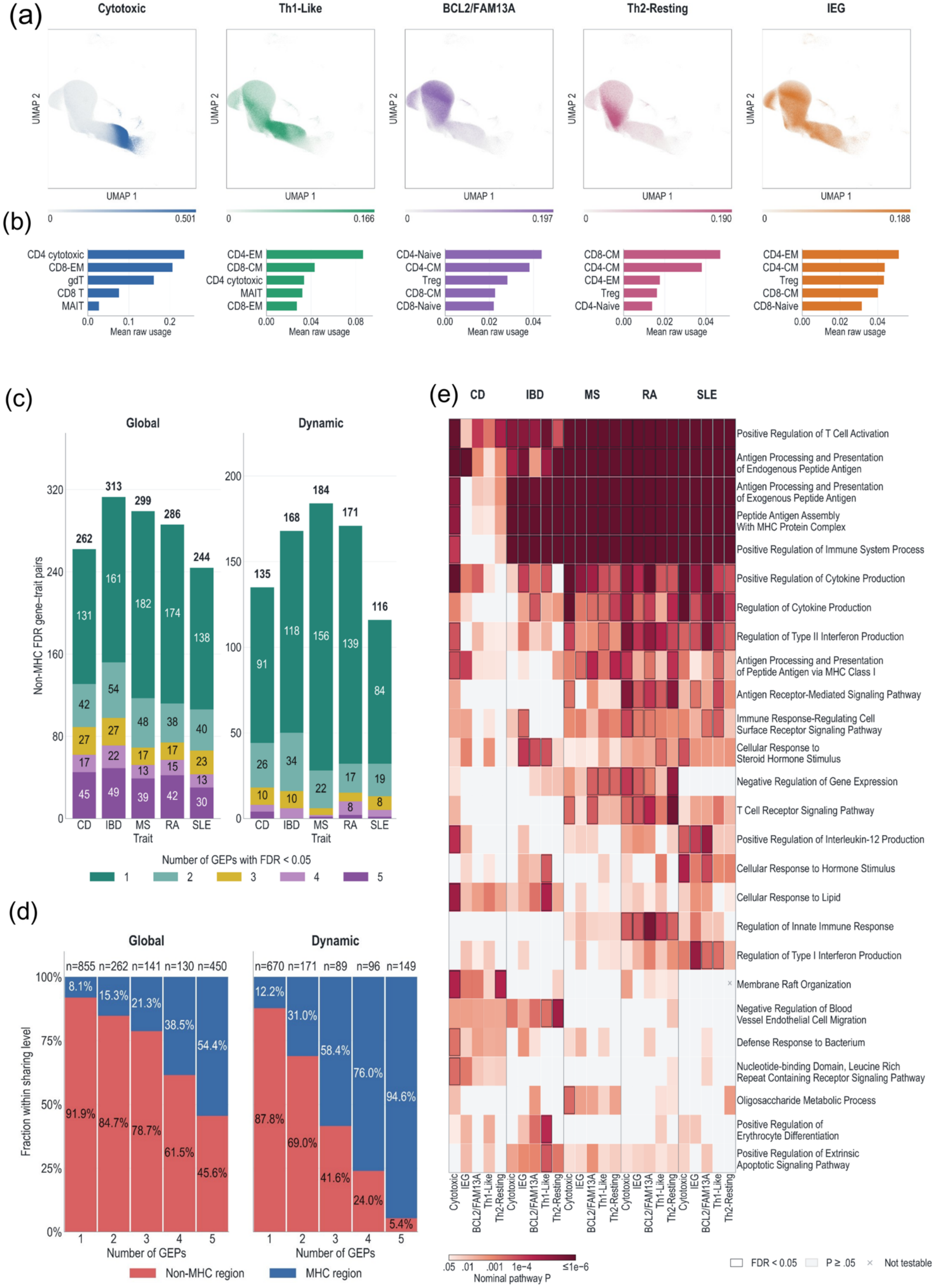
Gene-disease associations along T cell gene expression programs (GEPs). (a) OneK1K T-cell UMAP colored by GEP usage value. (b) Mean usage value in the five highest-usage cell types. (c) Number of significant (FDR < 0.05) non-MHC genes associated with each disease by GEP sharing level. (d) GO Biological Process over-representation analysis of globally associated genes (FDR < 0.05) in each disease– GEP combination, including MHC genes. For each disease, terms with nominal P < 0.05 in at least one GEP were grouped by gene-set overlap (Jaccard similarity ≥ 0.25). Up to eight representative terms were selected per disease and combined, yielding 26 unique terms. See Methods for detailed selection strategy. Colors indicate nominal hypergeometric P values, capped at P ≤ 10^−6^; black outlines indicate Benjamini–Hochberg FDR < 0.05, and gray indicates P ≥ 0.05. A cross (×) indicates a pathway not evaluated because fewer than 10 pathway genes were present in the corresponding tested-gene background. (e) Proportion of MHC and non-MHC genes by GEP sharing level; n denotes the number gene-trait pairs. CD, Crohn’s disease; IBD, inflammatory bowel disease; MS, multiple sclerosis; RA, rheumatoid arthritis; SLE, systemic lupus erythematosus.

For each of the five GEP-defined continuous states, we trained GReX prediction models with Fast-TWiST (compressed FLASSO) and conducted association analyses for five immune-mediated diseases: RA, SLE, CD, IBD and MS, yielding 25 disease-GEP combinations. AS and T1DM were excluded because their GWAS had the smallest effective sample sizes among the seven candidate diseases. Across the 25 disease-GEP pairs, Fast-TWiST identified 5,826 global gene-disease-GEP associations, and 3,092 dynamic associations with varying effects along GEPs (FDR < 0.05, Supplementary Table S1). These associations represent 1743 unique global genes and 1073 unique dynamic genes. For individual disease-GEP pairs, the number of dynamic genes ranged from 57 genes for CD in the IEG program to 194 genes for RA in the Cytotoxic program (Supplementary Table 1). Among genes outside of MHC and gene–disease pairs tested in all five GEPs, 56.0% (786/1,404) of globally associated 76.0% (588/774) of dynamically associated gene-disease pairs were GEP-specific (Figure 4c), while 205 and 8 pairs were shared across five GEPs, respectively. Including MHC genes substantially increased the proportion of program-shared genes, especially for dynamic tests (Supplementary Figure S6). From genes specific to one GEP to genes shared by 5 GEPs, the proportion of gene-disease pairs involving MHC genes increased from 8.1% to 54.4% for global associations and from 12.2% to 94.6% for dynamic (Figure 4d).

To characterize the biological processes represented by Fast-TWiST discoveries, we performed over-representation analysis (ORA) of Gene Ontology biological process terms separately for global and dynamic discoveries in each disease–GEP combination. Both analyses identified enrichment of antigen processing and presentation of endogenous peptide antigen (GO:0002483), regulation of cytokine production (GO:0001817), and positive regulation of T cell activation (GO:0050870) across multiple diseases and GEPs (Figure 4e; Supplementary Figure S7). Alongside these shared processes, dynamic were enriched in several unique pathways, including regulation of protein ubiquitination among IBD-associated genes in the IEG program (BH q = 0.0098) and epigenetic regulation of gene expression among MS-associated genes in the Th1-Like program (BH q = 0.011; Supplementary Figure S7). Excluding MHC-region genes substantially reduced the number of enriched GO terms. The number of terms reaching FDR<0.05 in at least one disease–GEP combination decreased from 96 to 8 for global discoveries and from 73 to 2 for dynamic discoveries (Supplementary Figures S8 and S9).

Nevertheless, the MHC-excluded analyses identified enrichment of negative regulation of T cell receptor signaling pathway among global RA-associated genes in the Cytotoxic, IEG, and BCL2/FAM13A programs (Supplementary Figure S8), and positive regulation of T cell activation among dynamic CD-associated genes in the Th1-Like program (Supplementary Figure S9).

### Cell-state-dependent regulatory effects near dynamic genes

We further examined whether the dynamic genes we discovered are supported by dynamic regulatory mechanisms using independent T-cell eQTL and single-cell multiome data. Nathan et al^15^ characterized eQTL effects across continuous states of more than 500,000 unstimulated memory T cells. Soskic et al^34^ identified cell activation-dependent eQTLs in naïve and memory CD4^+^ T cells. From Onek1k, we discovered 3,092 significant gene–disease–GEP associations, corresponding to 1,073 unique genes. Among the 1,073 dynamic, 180 overlapped state-dependent eGenes in Nathan et al. and 175 overlapped dynamic eGenes in Soskic et al., yielding 290 genes (27.0%) supported by at least one of the datasets (Supplementary Figure S10). To examine whether the regulatory SNPs for the dynamic genes are located in chromatin regions whose accessibility depends on cell states, we analyzed paired RNA-ATAC profiles from 5,942 T cells in a public PBMC multiome dataset from a single healthy donor (see Methods for details). For 719 out of 1,073 dynamic genes (67%), at least one ATAC-seq peak containing a Stage-1 predictor SNP or a strong LD proxy (R^2^>0.8) varied in accessibility along the same GEP in which the gene showed a dynamic association. These results support cell-state-dependent regulatory effects on these genes, which may be underlying dynamic associations with diseases.

Next, we turned our attention to two gene-disease pairs with dynamic associations. Fast-TWiST identified a dynamic ORMDL3–SLE association along the Cytotoxic program (q = 9.3 × 10^−6^), with the fitted association changing from negative at the lowest GEP rank (*β* = −1.57) to positive at the highest rank (*β* = 1.99, Figure 5a). rs2290400 lies at the 17q21 locus and is one of the SNPs in the Stage-1 prediction model for ORMDL3. It is in strong LD with rs2313430 (r^2^ = 0.914 with rs2290400), which lies within an ATAC peak showing increased accessibility with increased usage value of the Cytotoxic program (Figure 5b). rs2313430 was also in strong LD with the SLE GWAS variant rs8074437 (r^2^ = 0.852, Figure 5c).^29^

**Figure 5.**
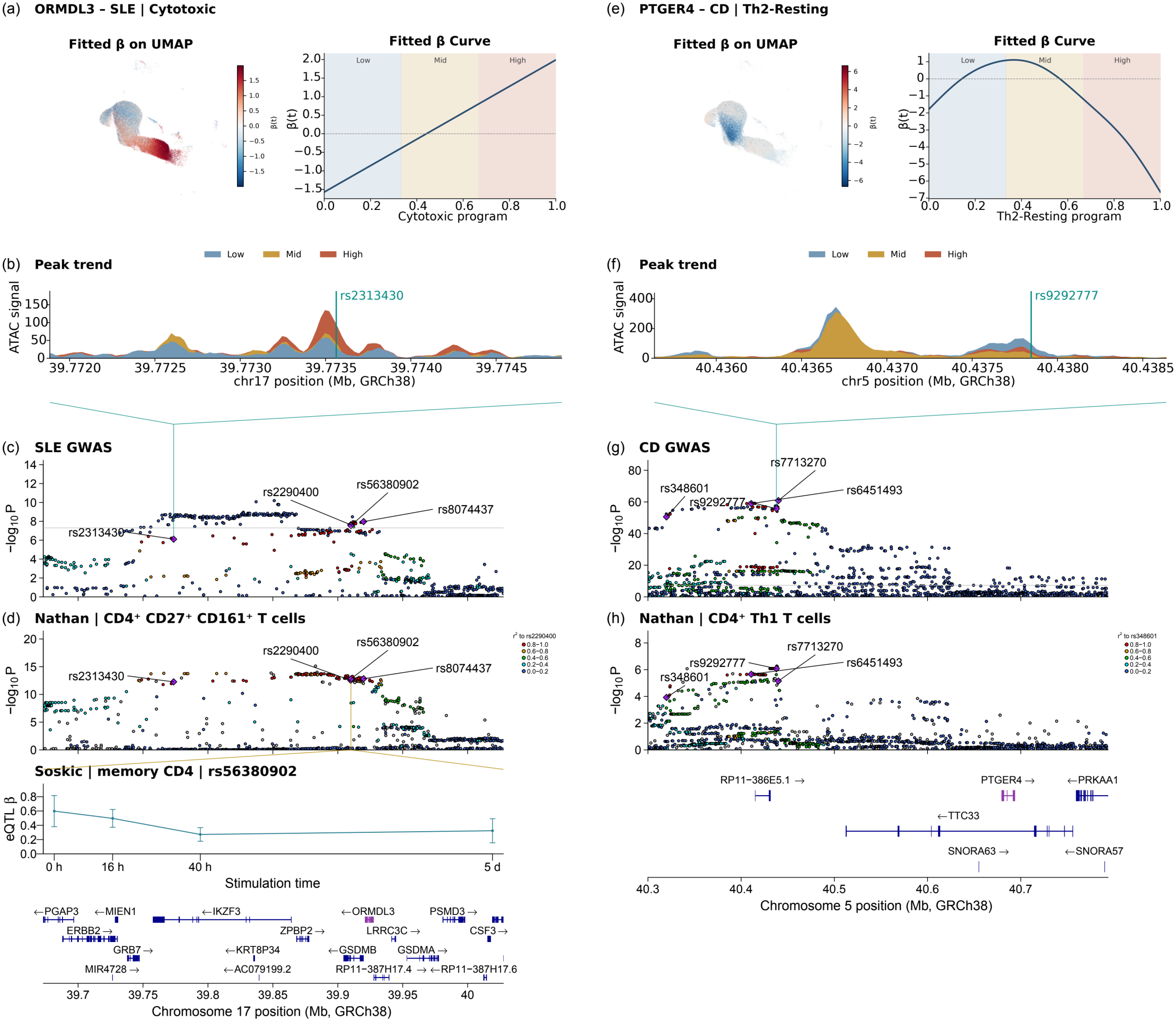
GEP-dependent regulatory effects near ORMDL3 and PTGER4. (a) Fitted Fast-TWiST association effects, *β*(*t*), across the Cytotoxic program for ORMDL3-SLE. The curve on the right shows *β*(*t*) versus cell GEP rank. Background colors indicate low, middle and high rank thirds. (b) Depth-normalized ATAC-seq signal around ORMDL3 in T cells. Cells were grouped into low, middle and high GEP-rank thirds in the dataset. (c) SLE GWAS local Manhattan plot near ORMDL3. (d) Local eQTL Manhattan plot near ORMDL3 for CD4^+^ CD27^+^ CD161^+^ T cells based on results reported in Nathan et al, and eQTL effects in memory CD4+ T cells by stimulation time based on results reported in Soskic et al. (e-h) Same plots for PTGER4 and CD in Th2-Resting, except that eQTL effects from Soskic et al were not included in (h). CD: Crohn’s disease. SLE: systemic lupus erythematosus.

Previous eQTL studies also discovered state-dependent genetic regulation of ORMDL3 at this locus. Nathan et al^15^ also studied ORMDL3 and reported significant interactions between the eQTL variant rs4065275 (r^2^ = 0.761/0.894 with rs2313430/rs8074437) and continuous T-cell states, including cytotoxicity, with the strongest eQTL effects observed in GZMB^+^ cytotoxic CD8^+^ T cells. Soskic et al^34^ reported another variant, rs56380902 (r^2^ = 0.996 with rs2290400, Figure 5d), associated with activation-dependent ORMDL3 regulation in memory CD4^+^ T cells. In addition, Perez et al^14^ reported colocalization between SLE GWAS signals and ORMDL3 eQTLs in CD8^+^ T cells (PP = 0.92). These findings further support context-dependent genetic regulation of ORMDL3 at a locus associated with SLE risk.

In another example, PTGER4 has a dynamic association with CD for Th2-resting program (q = 3.7 × 10^−10^): the fitted association changing from negative at the lowest GEP rank (*β* = −1.78) to positive at intermediate ranks (*β* = 0.61), before declining to negative at the highest rank (*β* = −6.65, Figure 5e). rs348601 lies at the 5p13.1 locus and is one of the SNPs in the Stage-1 prediction model for PTGER4. It is in strong LD with rs9292777 (r^2^ = 0.855), which lies within an ATAC peak showing higher accessibility in cells with low usage values of than those with medium and high usage values of Th2-Resting (Figure 5f). rs9292777 was also in strong LD with the CD GWAS variant rs7713270 (r^2^ = 0.996, Figure 5g).^30^

Nathan et al^15^ found that the effect of the PTGER4 eQTL variant rs6451493 (r^2^ = 0.870/0.975 with rs348601/rs9292777) on gene expression varied significantly across memory T-cell states overall (P = 3.0 × 10^−13^), including along a Th2-related transcriptional state (P = 2.9 × 10^−7^).

They also found that rs9292777 was associated with PTGER4 expression in CD4^+^ Th1 cells (cis-SNP Bonferroni-adjusted P = 0.0064, Figure 5h). In addition, another study^35^ reported associations between CD susceptibility variants and PTGER4 expression. These findings further support context-dependent genetic regulation of PTGER4 at a locus associated with CD risk.

## Discussion

In this paper, we present Fast-TWiST, a scalable method for TWAS at cell-state resolution. Fast-TWiST extends our previous method^36^ to accelerate the training of gene expression prediction models, running >10 times faster than TWiST while maintaining comparable statistical power. The improved speed enabled its application to raw single-cell data in the OneK1K study without meta-cell aggregation. The genes we identified had a high replication rate in an independent dataset. A new application using GEPs to characterize cell states, identified hundreds of dynamic genes along interpretable dimensions of cell state. These findings were supported by dynamic regulatory effects in independent eQTL and single-cell multiome datasets.

Fast-TWiST includes two methods, compressed FLASSO and FPCA, and a CCT to combine their P-values. Because compressed FLASSO fits the same model as TWiST, they had similar statistical performance. However, due to its faster speed, compressed FLASSO could be applied to single cells without meta-cell aggregation, it led to stronger association signals than TWiST applied to meta-cells (Figure 3). The FPCA, on its own, appears to be less powerful than compressed FLASSO in many scenarios for the global and dynamic tests, and have no power for the nonlinear test. It appears to perform better with less noisy data, surpassing the power of compressed FLASSO in a number of cases (Supplementary Figure S1). A possible explanation is that FPCA tends to project gene expression onto a small number of FPCs by design, and in some cases leads to very few basis functions and predictor SNPs. This dimension reduction is useful for data representation, but for identifying complex curves in Stage 2, does not provide enough number of dimensions for the matrix algebra. For example, conducting a nonlinear test requires at least 3 FPCs and 3 predictor SNPs with non-zero effects. If FPCA projects all signals to two dimensions, the nonlinear test cannot be performed. Therefore, we do not recommend using FPCA on its own. However, the CCT, which combines compressed FLASSO and FPCA, led to higher power than compressed FLASSO in some scenarios. This demonstrates that FPCA may contribute signal to the test by capturing complementary patterns to those captured by compressed FLASSO.

Another novel aspect of the paper is the GEP-based TWAS analysis. In our previous study^36^, we conducted cell-state-resolved TWAS for CD4^+^ T cells, CD8^+^ T cells and B cells, and used pseudotime to capture the transition from naïve to memory cells. However, a one-dimensional pseudotime cannot capture the full heterogeneity of T cells, which comprises many subgroups with distinct functions. We avoided this complexity in the earlier study by conducting analyses within naïve and central memory cells and removing subgroups that do not fit the framework, such as regulatory T cells and gamma-delta T cells. By using GEPs in this study, we captured more complex, biologically interpretable cell states and identified novel dynamic genes.

Transition along these cell states were aligned with the expected change in cell type proportion among discrete cell types (Supplementary Figure S5). Among the five GEPs we investigated, we identified thousands of gene-disease associations whose effect vary dynamically along GEP usage values. We found that genes that are shared by multiple GEPs are highly concentrated in the MHC region, and non-MHC genes tend to be GEP-specific. We also conducted in-depth follow-up analyses for two genes. For both, converging evidence from multiple independent sources supported cell-state-dependent regulatory effects that may underlie their cell-state-dependent disease associations. These genes are candidate targets for precision therapies directed at specific cell subpopulations, which could reduce off-target side effects.

This study has a few limitations. First, Fast-TWiST improves computational speed, but only minimally improves statistical power. Future work will be devoted to further improve the method across statistical power, estimation efficiency, and other evaluation metrics. Second, since Fast-TWiST does not change the model of TWiST, it is still designed for one-dimensional cell-state variables. Therefore, it cannot handle more sophisticated situations, such as multiple branches in a differentiation trajectory. Third, although we used several single-cell eQTL datasets in this paper, it is far from an exhaustive list. Integrating other datasets can capture a wider range of cell states, but harmonizing them raises challenging heterogeneity issues, such as study population, experimental protocol, and processing pipelines. Addressing these issues is beyond the scope of this paper and is a direction for our future work.

In conclusion, Fast-TWiST is a scalable and powerful method for TWAS at cell-state resolution. With the rapid growth of single-cell eQTL data, Fast-TWiST can help uncover new cell-state-dependent gene-disease associations and prioritize targets for precision interventions.

## Methods

### Model setup

Built on our previous work^24^, we developed Fast-TWiST, a scalable method for TWAS at cell-state resolution. In a single-cell eQTL study, we assume that genotypes and single-cell gene expression are measured for *I* individuals. For a gene of interest, denote by *x*_*ij*_ the unique molecular identifier (UMI) count for cell *j* (*j* = 1, …, *J*_*i*_) from individual *i* (*i* = 1, …, *I*); *N* = ∑_*i*_ *J*_*i*_ denotes the total number of cells. Each cell has a continuous cell state variable *t*_*ij*_ (e.g., pseudotime or other cell embeddings), library size α_*ij*_, and a vector of individual- and cell-level covariates (e.g., age, sex, expression and genotype principal components) denoted by ***z***_*ij*_. Throughout, *t*_*ij*_ is the rank-transformed cell state,

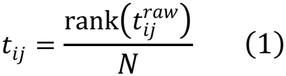

where 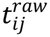 is the raw cell state value, e.g., pseudotime produced by software such as Slingshot^37^ or TSCAN^38^ or the usage value of a GEP, so that *t*_*ij*_ is approximately uniformly distributed on [0,1]. Each cell from the same individual has the same value for individual-level covariates. Denote by *g*_*ik*_ the genotype of individual *i* at cis-SNP *k* (*k* = 1, …, *K*), standardized to mean 0 and variance 1, and further define ***g***_*i*_ = (*g*_*i*1_, …, *g*_*ik*_)^⊤^.

Following TWiST, we model single-cell gene expression counts using a Poisson distribution in Stage 1, and link mean expression to genotype through a function-on-scalar regression along pseudotime:

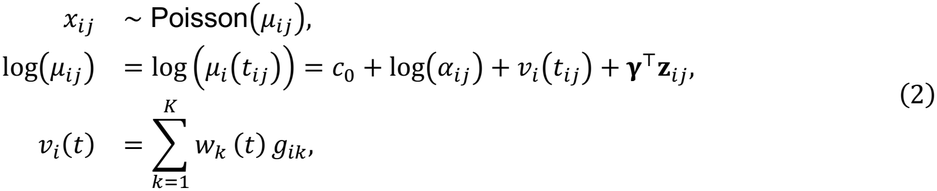

where *v*_*i*_(*t*) is the genetically regulated expression (GReX), *c*_0_ is an intercept, and γ is a vector of covariate effects. Each SNP’s regulatory effects on GReX, *w*_*k*_(*t*), is allowed to vary smoothly across cell state *t* rather than being a single scalar effect. In the original TWiST, *w*_*k*_(*t*)’s was parameterized as B-spline functions and estimated using group lasso implemented in the grpreg package in R^39^, which was computationally intensive when the number of cells is large. Fast-TWiST adopts several strategies to accelerate the estimation of *w*_*k*_(*t*), described below.

The estimation of *w*_*k*_(*t*) requires many iterations of numerical optimization across multiple cross-validation folds. To avoid re-estimating nuisance parameters (*c*_0_, γ) at every iteration, we first regress out the covariates by fitting a covariate-only Poisson regression by maximum likelihood,

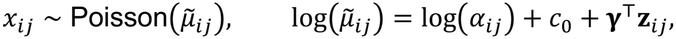

using standard R function glm. The fitted linear predictor, 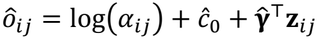, is then treated as a known offset, and the GReX trajectory is estimated from Model#(2)#(2) (2) by fitting log 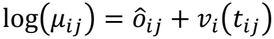 with 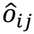 held fixed. The intercept (*c*_0_) and covariate effects (γ) are thus estimated only once and used for all downstream iterations. We propose two methods for estimating *v*_*i*_(*t*), described below (Stage 1a and 1b).

### Stage 1a: Compressed FLASSO

As in TWiST^24^, each SNP effect is modeled as a linear combination of a common set of cubic B-spline basis functions *ϕ*(*t*) = (ϕ_1_(*t*), …, *ϕ*_*M*_(*t*))^’^,

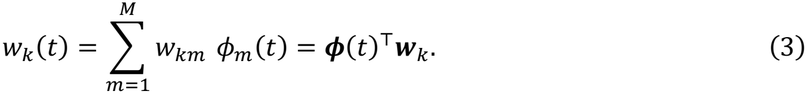

This specification converts the problem to a Poisson regression with *KM* predictors. The *M* predictors for SNP *k* form an *N* × *M* design block **G**_*k*_, whose row for cell *j* of individual *i* is *g*_*ik*_ *ϕ*(*t*_*ij*_)^⊤^. Following grpreg, each design block is centered and transformed, 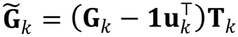, before model fitting so that 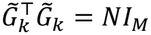. Here **u**_*k*_ is the vector of column means of **G**_*k*_, and **T**_*k*_ is an *M* × *M* matrix, obtained through eigendecomposition, that scales and orthogonalizes the centered block (Supplementary Methods S1). The coefficients 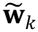 of the transformed blocks are estimated by minimizing the average negative Poisson log-likelihood plus the group elastic-net penalty:

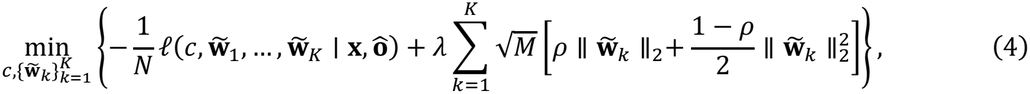

where *l*(·) is the Poisson log-likelihood with mean exp 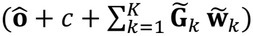, and *c* is an unpenalized intercept. The penalty term aims to introduce sparsity at the SNP level, shrinking all *M* of its coefficients together. Here *ρ* is a tuning parameter to balance between L1 and L2 group penalties. As in TWiST, we set *ρ* = 0.5 in all analyses. The regularization parameter λ is selected by cross-validation. The coefficients in the original spline basis are then recovered as 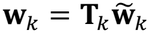.

#### Why a direct fit is slow

We previously fitted the model using grpreg,^39^ which constructs the full *N* × *KM* cell-level design matrix. This matrix is large: *N* often exceeds 1 million, and *K* is typically in the hundreds when restricted to HapMap 3 SNPs and thousands otherwise. More importantly, it is used repeatedly during the model fitting. grpreg minimizes (4) by coordinate descent^39^: each iteration cycles through the *K* SNPs, and for each SNP it (i) computes the cross-product 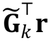 between the SNP’s design block and the current working residuals **r** = (*r*_*ij*_), where *r*_*ij*_ = (*x*_*ij*_ − *µ*_*ij*_)/*max*_*i,j*_*µ*_*ij*_ (Supplementary Methods S1), and (ii) after updating 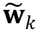 updates the residuals of all cells. Both steps sum over all *N* cells, which is computationally intensive when *N* is large. This cost recurs in every iteration, for every value of λ, in every cross-validation fold, and for every gene.

#### Key idea: genotypes are constant within an individual

All cells from individual *i* share the same genotype *g*_*ik*_, so the genotype can be moved outside the sum over that individual’s cells:

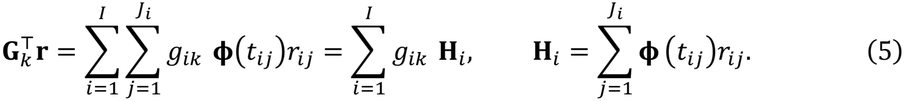

It follows that 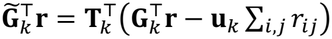, where the last term is zero after the intercept update (Supplementary Methods S1). The residual summaries **H**_*i*_ are *M*-vectors that do not depend on the SNP *k*. They are computed in a single pass over the cells and shared by all *K* SNPs, so the cross-product for each SNP becomes a sum over *I* individuals instead of *N* cells. The number of individuals (*I*) is typically orders of magnitude smaller than *N*. After each update of 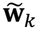, **H**_*i*_ must also be updated. Although **H**_*i*_ is defined as a sum over cell in (5), it can be updated at the donor level:

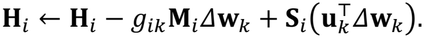

See Supplementary Methods S1 for details. Here 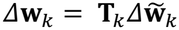 is the change in **w**_*k*_ in this iteration, and **S**_*i*_ and **M**_*i*_ are two individual-level summary statistics:

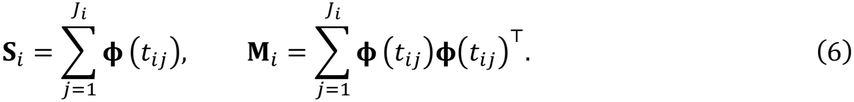

These statistics do not depend on the model parameters, so they are computed before the iterations begin and held fixed across iterations. This allows **H**_*i*_ to be updated rapidly.

### Stage 1b: FPCA

Principal component analysis (PCA) is a dimension reduction technique that projects multivariate data onto a small number of directions capturing the largest variation. Functional principal component analysis (FPCA) extends this idea to curves, summarizing variation across individual curves with a small number of shared patterns. Each pattern is represented by a function called a functional principal component (FPC), and each individual curve can be approximated by a linear combination of FPCs. In the TWAS setting, the GReX trajectory can likewise be approximated by a linear combination of functions representing distinct expression patterns. For example, one pattern may describe differences in overall expression level throughout the cell-state trajectory, whereas another may describe higher expression in early states and lower expression in late states. These patterns are learned from the data rather than specified in advance. Since gene expression data are non-Gaussian, conventional FPCA methods^40^ are not directly applicable. We therefore extended Fast-GFPCA^41^ to estimate the FPCs (Steps 1 and 2). The original Fast-GFPCA assumes that all individuals are observed at the same set of time points. Our extension removes this assumption and allows each individual to have their own set of time points. After estimating the FPCs, we regress the FPC scores on genotypes to construct GreX prediction models (Step 3).

**Step 1**. First, we estimate individual-level expression curves from single-cell counts using Poisson mixed-effects models. We partition the pseudotime range [0,1] into bins of equal width *h*, yielding *R* = 1/*h* bins with approximately equal number of cells. Any bin in which all expression counts are zero is merged with an adjacent bin.

Let *r*(*t*) denote the resulting bin containing pseudotime *t*. Using the offset 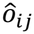 from the covariate-only model in Model Setup, we fit a Poisson mixed-effects model separately within each bin,

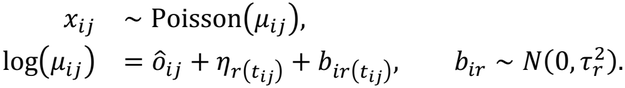

Here, η_*r*_ is the bin-specific intercept, *b*_*ir*_ is the random effect for individual *i* in bin *r*, and 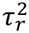 is the between-individual variance within that bin. The estimated random effect 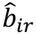 summarizes the individual’s deviation on the log-expression scale after adjusting for library size and covariates. We treat 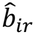 as an observation of an individual-specific trajectory *b*_*i*_(*t*) evaluated at the midpoint at bin *r*.

**Step 2**. Second, we infer functional PCs from individual-specific trajectories. We estimate the mean curve 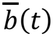 and the covariance function *C*(*s, t*) = Cov{*b*_*i*_(*s*), *b*_*i*_(*t*)}, which are smooth across time *s* and *t*, across individuals using fpca.sc from the refund package^42–44^. The smooth eigenfunctions *φ*_*l*_(*t*) of this covariance, also inferred by fpca.sc, describe patterns of deviation from the mean curve shared across individuals. They are orthonormal and ordered by their eigenvalues *v*_1_ ≥ *v*_2_ ≥ … ≥ 0, so that earlier components explain more between-individual variation. Retaining the first *L* components gives

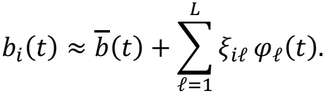

The score 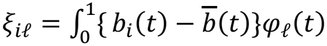 *dt* describes the magnitude and direction of individual *i*’s deviation along component *l* and is an output of fpca.sc. We choose the smallest *L* for which the cumulative proportion of variance explained, 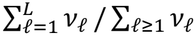, reaches a prespecified threshold. The mean, covariance, eigenfunctions, and scores are estimated separately for each gene.

**Step 3**. Third, we train models to predict the scores from genotype. Since *φ*_*l*_(*t*)’s are shared across individuals, the scores determine individual-to-individual variation in gene expression. We regress the estimated scores on cis-SNP genotypes using the linear model

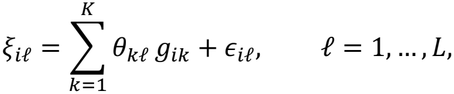

where θ_*kl*_ is the effect of SNP *k* on component *l* and ϵ_*il*_ is the error term. The genotype coefficients are estimated by elastic-net regression using glmnet^45^ with elastic-net mixing parameter 0.1 (the weight of the L1 penalty, with 0.9 on the L2 penalty), with the regularization parameter selected by cross-validation across individuals. The predicted GreX 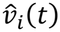 is then

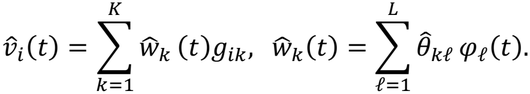

The shared mean curve 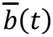 is not included in GReX because it is shared across individuals and does not represent genotype effects. Compressed FLASSO and FPCA therefore estimate the same quantity, a genotype-predicted expression trajectory, through different Stage 1 models. Either model can be used in Stage 2.

### Stage 2. Estimating the effect of gene expression on a trait

We use the original TWiST Stage 2 model for both Stage 1 methods; For FPCA, the eigenfunctions *φ*_*l*_(*t*) take the place of the B-spline basis in the Stage 2 computations (Supplementary Methods S2). Denote by *y*_*a*_ the trait value for individual *a* in the GWAS sample, which may not overlap the individuals used for Stage 1 training. Assume that *y*_*a*_’s are adjusted for covariates, and standardized to have mean 0 and unit variance. Using models trained in Stage 1, we impute the GReX curve as 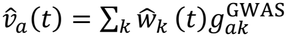, where 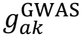 is the genotype value of SNP *k* of individual *a* in the GWAS sample. We model *y*_*a*_ using scalar-on-function regression

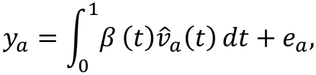

where *e*_*a*_ is a mean-zero residual and *β*(*t*) describes the association between GReX and the trait along pseudotime. We represent this effect as

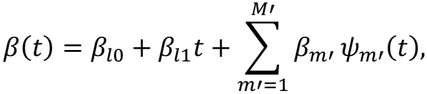

where *β*_*l*0_ and *β*_*l*1_ are the coefficients of the constant and linear terms, and *ψ*_*m’*_(*t*) are the B-spline basis functions generated by bs(…, intercept = TRUE) in R package splines, excluding the first and last B-spline basis functions to ensure linear independence. Smoothness is controlled through the random-effects representation 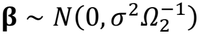, where *β* = (*β*_1_, …, *β*_*M*’_) ^⊤^ and 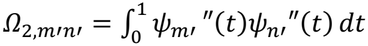. Setting *σ*^2^ = 0 removes this component, leaving a linear effect curve (Supplementary Methods S3).

The model can be fitted using individual-level data or GWAS summary statistics and a linkage-disequilibrium (LD) reference panel (Supplementary Methods S2–S4). We test three nested null hypotheses:

- **Global test:** no association at any pseudotime value, *H*_01_: *β*(*t*) = 0 for all *t*, corresponding to *β*_*l*0_ = *β*_*l*1_ = 0 and *σ*^2^ = 0.
- **Dynamic test:** no change in the effect along pseudotime, *H*_02_: *β*(*t*) = *β*_*l*0_ for all *t*, corresponding to *β*_*l*1_ = 0 and *σ*^2^ = 0.
- **Nonlinear test:** no departure from a linear effect, *H*_03_: *β*(*t*) = *β*_*l*0_ + *β*_*l*1_*t*, corresponding to *σ*^2^ = 0.

We use likelihood-ratio tests with the mixture reference distributions as in the TWiST paper.

### Combining compressed FLASSO and FPCA: Cauchy combination test

The compressed FLASSO (Stage 1a) and FPCA (Stage 1b) estimators of *v*_*i*_(*t*) may capture different patterns of pseudotemporal variation of gene expression. To leverage the strengths of both, we apply the Stage 2 association tests to each estimator and combine the resulting P-values using the Cauchy combination test (CCT). Let *P*_*C*_ and *P*_*F*_ denote the P-values from the same Stage 2 test (global or dynamic) for compressed FLASSO and FPCA, respectively. The CCT statistic and P-value are calculated as

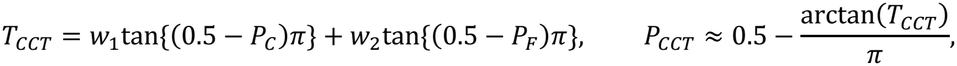

with equal weights *w*_1_ = *w*_2_ = 0.5. This approximation remains accurate under arbitrary dependence between *P*_*C*_ and *P*_*F*_, particularly at small significance levels^26^. This property is important because both P-values are derived from the same underlying single-cell data and their dependence structure is difficult to quantify. We do not combine nonlinear P-values because the null distribution of its test statistic has a point mass at zero, causing the approximation to break down (Supplementary Methods S4).

### Simulation studies

We followed the simulation design of the TWiST study. Gene expression was simulated using real chromosome 1 genotype data from 1,033 individuals in the OneK1K cohort. Each combination of simulation parameters included 200 randomly selected genes with at least 300 cis-SNPs within 500 kb of the transcription start site. A total of 50,000 cells were randomly assigned to the 1,033 individuals, with the underlying time variable *t* sampled from Uniform(0,1).

For each gene, 5, 10, or 20 cis-SNPs were randomly selected as causal variants. Their effects on expression across *t* were represented using cubic B-splines with 1, 3, or 5 equally spaced internal knots. The knot locations were 0.5 for one knot; 0.25, 0.5, and 0.75 for three knots; and approximately 0.16, 0.33, 0.5, 0.67, and 0.83 for five knots.

For each causal SNP *k*, the coefficient corresponding to basis function *m* was generated as *w*_*km*_ = *a*_*k*_ + *d*_*km*_, where *a*_*k*_ ~ *N*(0,0.01) was a shared SNP-level component and *d*_*km*_ ~ *N*(0,0.01) was an independent basis-specific component. The SNP-effect curve was *w*_*k*_(*t*) = ∑_*m*_ *w*_*km*_ *ϕ*_*m*_(*t*), giving the GReX trajectory *v*_*i*_(*t*) = ∑_*k*_ *w*_*k*_ (*t*)*g*_*ik*_ for individual *i*. All coefficients for noncausal SNPs were set to zero.

Expression counts were generated from Poisson distributions with mean exp{*v*_*i*_(*t*_*ij*_) + *t*_*ij*_ + ϵ_*ij*_}. The term *t*_*ij*_ represented an effect of time *t* on expression that was independent of genotype. In the 10x-noise setting, the residual term ϵ_*ij*_ was drawn independently from 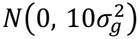 in the no-noise setting, ϵ = 0. Here 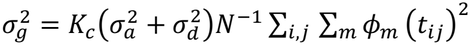 approximates the variance of the genetic component across cells, with *K*_*c*_ being the number of casual SNPs, *N* the number of cells, and 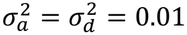 the variances of *a* and *d*. This gives a signal-to-noise ratio of approximately 0.1 on the log-mean scale which reflects high level of variation at cell level. All cells had the same library size, which was therefore omitted from the expression model.

The next step was to generate GWAS summary statistics for a complex trait. As in the original TWiST simulation, we generated the estimated marginal SNP effects directly from

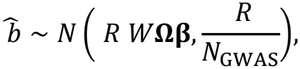

where *R* is the LD matrix calculated from the OneK1K genotype data, *W* = {*w*_*km*_} a *K* × *M* matrix containing the true SNP-effect coefficients, *β* = (*β*_1_, …, *β*_*M*_)^⊤^ are the coefficients of the true effect curve *β*(*t*) = ∑_*m*_ *β*_*m*_ *ϕ*_*m*_(*t*), represented in the same B-spline basis as the SNP effects, Ω is the *M* × *M* matrix with entries 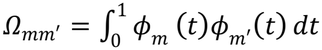, and *N* = 100,000.

We generated *β* = (*β*_1_, …, *β*_*mid*−1_, *β*_*mid*_, *β*_*mid* + 1_, …, *β*_*M*_) under four scenarios:

- Null: *β*_1_ = … = *β*_*M*_ = 0;
- Constant: *β*_1_ = … = *β*_*M*_ = 0.05;
- Unimodal: 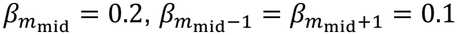, and *β*_*m*_ = 0 for all other *m*;
- Reverse: 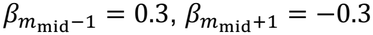, and *β*_*m*_ = 0 for all other *m*.

To analyze the simulated data, empirical pseudotime was estimated as the first principal component of the log(1 + *x*)-transformed expression matrix. Its direction was oriented to agree with the simulated trajectory, i.e., if the empirical pseudotime has a negative correlation with *t*, we flipped the sign of the empirical pseudotime, making the correlation positive. The values were then ranked and divided by the number of cells, giving an approximately uniform distribution between 0 and 1. TWiST, compressed FLASSO, and FPCA used the same empirical pseudotime values for Stage 1 training.

We applied TWiST and compressed FLASSO using cubic B-splines with three internal knots at 0.25, 0.5, and 0.75. Both methods selected the penalty tuning parameter from 20 candidate values using five-fold cross-validation with the same fold assignments. For FPCA, cells were ordered by empirical pseudotime and divided into approximately 50 bins, corresponding to a target bin width of 0.02. Library-size offsets and covariate terms were omitted because library size was constant and no additional covariates were simulated. After applying FPCA, we retained the top functional principal components that together explained at least 99% of the variation in these trajectories. Each component score was regressed on cis-SNP genotypes using elastic net with mixing parameter 0.1 and a penalty parameter selected by cross-validation. The fitted genotype effects were projected through the estimated eigenfunctions to obtain the SNP-effect curves.

For Stage 2, we used cubic B-splines with 19 internal knots at 0.05,0.10, …,0.90,0.95 to estimate *β*(*t*). TWiST, compressed FLASSO, and FPCA were evaluated using the same global, dynamic, and nonlinear tests. We combined the global or dynamic P-values from compressed FLASSO and FPCA using the Cauchy combination test with equal weights. Nonlinear P-values were not included in the combination.

We also compared the three functional methods with pseudobulk lasso and elastic net. For the global test, all cells from the same individual were aggregated into one pseudobulk sample. The average count across cells was calculated for each individual and log-transformed. For the dynamic test, cells were divided at empirical pseudotime 0.5 and aggregated separately into early (pseudotime≤0.5) and late (pseudotime>0.5) pseudobulk samples. Prediction models were trained separately for the two samples, and equality of their estimated trait effects was tested.

For each combination of the number of causal SNPs and the true number of internal knots, we repeated the simulation three times. This produced 27 simulated expression datasets and 600 genes for each combination of the number of causal SNPs and internal knots, for a total of 5,400 gene-level simulations in each noise setting.

### The OneK1K cohort

The OneK1K study^12^ recruited 1,104 individuals of predominantly Northern European ancestry. The published single-cell dataset comprises approximately 1.27 million peripheral blood mononuclear cells from 982 donors. Genotype data were obtained from the Gene Expression Omnibus (GEO; GSE196830), and processed single-cell gene-expression data and cell annotations were obtained from the Human Cell Atlas/CELLxGENE collection.

### Genotype data processing and quality control

Genotype calls were exported from GenomeStudio in PLINK format. Downstream quality control was performed using PLINK. Variant-level quality control retained biallelic autosomal SNPs with a call rate > 0.97, a minor allele frequency (MAF) >0.01, and a Hardy–Weinberg equilibrium exact-test P-value >1×10^−6^. Donors with a genotype call rate <0.97 or genome-wide autosomal heterozygosity rate more than three standard deviations from the sample mean were excluded. Pairwise genetic relatedness was estimated using GCTA,^46^ and one donor from each pair with estimated relatedness greater than 0.125 was removed. Following allele harmonization and pre-imputation quality control, 484,199 autosomal SNPs from 1,034 donors were retained.

Each autosomal chromosome was imputed separately through the Michigan Imputation Server v2.0.6^47^ using Eagle^47^ and Minimac v4.1.6, with the Haplotype Reference Consortium (HRC) r1.1 as the reference panel. The combined imputation output contained 39,117,105 variants across 1,034 donors. For downstream analyses, HapMap3 SNPs^48^ with imputation R^2^ greater than 0.8 and MAF greater than 0.01 were retained, yielding 1,173,026 SNPs.

To perform principal component analysis (PCA), we removed variants within predefined high-linkage-disequilibrium (LD) regions listed by the Center for Statistical Genetics at the University of Michigan (https://genome.sph.umich.edu/wiki/Regions_of_high_linkage_disequilibrium_(LD)). We further performed LD pruning to obtain a set of independent SNPs with r^2^<0.2. Genotype principal components were calculated from remaining SNPs. ()

### scRNA-seq processing and trajectory inference

Trajectory inference was conducted for each of the following cell types separately: CD4^+^ T cells (naïve and central memory), CD8^+^ T cells (naïve and central memory), and B cells (naïve, transitional, and memory). The gene expression of each cell type was normalized separately using SCTransform^49^, with 2,000 highly variable genes and the percentage of mitochondrial transcripts regressed out. PCA on gene expression data was performed, followed by Harmony correction^50^ for pool-associated batch effects using the first 50 principal components. UMAP embeddings were generated from the first 10 Harmony dimensions.

Slingshot^37^was fitted to the existing first 10 Harmony dimensions separately for each cell type. The inferred pseudotime was oriented from naïve to memory cells using the cell annotations. Pseudotime was then rank-transformed across the cells of each cell type as in (1), with tied values assigned their average rank. For model training, cells were matched to the available genotype data, retaining 547,509 CD4^+^ T cells, 68,889 CD8^+^ T cells, and 125,746 B cells from 980 donors in each cell type. Gene IDs in the input expression matrices were matched to the reference gene annotation to obtain chromosome locations and transcription start sites (TSSs). We retained autosomal genes with a matched TSS annotation and at least one available cis-SNP within 500 kb of the TSS in the genotype data. Genes without a matching annotation or any available cis-SNPs were excluded from model training. This yielded 12,920, 13,130, and 13,105 candidate genes for CD4^+^ T cells, CD8^+^ T cells, and B cells, respectively. All 980 genotype-matched donors were used for final model training. Fast-TWiST was applied to train gene expression prediction models using raw UMI counts, with age, sex, 10 expression principal components, and 10 genotype principal components as covariates, and an log library size as an offset.

### OneK1K association analysis

After training the Stage-1 model for CD4^+^ T cells, CD8^+^ T cells, and B cells, we retained for Stage-2 analysis only genes with at least three SNPs with non-zero coefficients, which we refer to as *Stage-1 valid* genes. Therefore, compressed FLASSO and FPCA each has a separate set of Stage-1 valid genes. We retained only Stage-1 valid genes for Stage-2 analysis, and the other genes did not have a Stage-2 P-value. For a gene to have a CCT P-value, it must be Stage-1 valid for both compressed FLASSO and FPCA. Prediction weights and GWAS summary statistics were aligned to the Phase 3 European LD reference panel from the 1000 Genomes Project. For all seven autoimmune diseases, we used the same GWAS summary statistics as in the original TWiST study. For each cell type, disease, and test, the performance of the methods were compared on a common gene set comprised of genes with P-values from all methods in the comparison. The QQ plots pooled raw P-values from these common sets across the seven diseases within each cell type (Figure 3b,c). Benjamini–Hochberg adjustment was applied separately for each method, cell type, disease, and test. Cis-SNPs were defined as those that are within 500kb upstream or downstream the transcription start site (TSS).

### The Perez et al. cohort: data processing and quality control

Genotype data were obtained from dbGaP accession phs002812.v1.p1^14^. The dataset comprised two cohorts (UCSF, N=209; ImmVar, N=49) sequenced using two genotyping platforms, which were processed separately. SNPs that were missing in more than 1% of the donors or with MAF≤ 1% were excluded. Samples with >1.5% missing genotypes were removed. Samples with autosomal heterozygosity rate greater than three standard deviations above the cohort mean were also removed. The resulting pre-imputation datasets contained 585,454 SNPs from 204 UCSF samples and 610,590 SNPs from 47 ImmVar samples.

Chromosome-specific VCF files were prepared in hg19 coordinates and submitted to the TOPMed Imputation Server using the TOPMed r3 reference panel^51^ and Eagle for haplotype phasing. Because the TOPMed r3 reference panel is based on GRCh38, the server converted hg19 input coordinates to GRCh38 during imputation. Imputed variants were retained if they were present in both platform-specific outputs, had imputation R^2^ greater than 0.3 in both cohorts, and were included in the HapMap3 reference set. Variants were harmonized across two cohorts to ensure they had the same chromosome, position, reference allele, and alternative allele before the two cohorts were merged.

The merged dataset was restricted to donors of European ancestry, and variants with MAF of 5% or lower were removed. This produced 940,371 SNPs from 146 donors in GRCh38 coordinates. Variants were subsequently converted to GRCh37/hg19 coordinates for compatibility with the downstream TWAS reference, retaining 939,986 SNPs from the same donors.

For the single-cell gene expression data, cells annotated as B_naive or B_mem in the original Perez et al. dataset were retained. After intersecting with the genotype data, the initial dataset contained 76,148 cells from 146 donors; After merging with the genotype data, 75,215 cells from 144 donors remained. Pseudotime was inferred using the same approach as the OneK1K data. We then removed genes with zero expression in >99.9% of B cells, retaining 13,025 genes.

### Validation in the Perez et al. cohort

Validation of the genes detected in OneK1K using our cell-state-based method required the pseudotime to represent a comparable transition of cell states in the discovery and validation cohorts. The cell type annotations provided by both datasets included B cells into naïve and memory cells. Since Perez et al did not include transitional B cells, we removed transitional B cells from OneK1K and reran Fast-TWiST analysis. This allows us to construct pseudotime trajectories using comparable cell populations in both datasets (Supplementary Figure S2). In addition, the CD4 and CD8 annotations differed substantially between OneK1K and Perez in subtype definition and granularity. Therefore, these cell types were not included in the validation. Fast-TWiST Stage-1 models were trained in the Perez B-cell data, independent of the models trained for OneK1K, using the same modeling framework. Age, sex, 10 expression PCs, and 5 genotype PCs were included as covariates. Only Stage-1 valid genes, i.e., those that have non-zero coefficients for at least 3 SNPs, entered Stage-1 analysis and received a P-value.

We performed Stage-2 Fast-TWiST analysis using GWAS summary statistics that are independent of those used for the OneK1K analyses. We used publicly available summary statistics for AS (Phe_715_2), IBD (Phe_555), MS (Phe_335), and RA (Phe_714_1) from the European-ancestry samples of the Million Veteran Program (MVP),^52^ as well as those for CD, T1DM and SLE from FinnGen^53^.

For each gene identified in the OneK1K analysis (FDR<0.05 for each disease-method-cell type combination), we checked whether it was significant in the Fast-TWiST analysis of Perez et al and the GWAS summary statistics listed above. Replication rates were computed with the number of genes that were significant in OneK1K and received a Stage-2 P-value in Perez et al as the denominator. Strict replication required the corresponding Perez association for the same gene, disease, and Fast-TWiST test to have FDR below 0.05. Nominal replication was defined as Perez raw P-value below 0.05. Global and dynamic associations were analyzed separately throughout. Genes were classified as belonging to the extended MHC when their annotated transcription start site (TSS) was on chromosome 6 between 25 and 35 Mb in GRCh37. MHC-stratified summaries were generated after cohort-specific FDR calculations; P and FDR values were not recalculated after MHC exclusion.

### Conventional FUSION cross-reference comparison

Conventional TWAS models were evaluated in subtypes of B cells using the following pipeline for both OneK1K and Perez et al. We first divided B cells by cell type annotations provided by the original study: naïve B cells and memory B cells. As an alternative, we also partition the B cells by pseudotime values: early (pseudotime<0.5) and late (pseudotime≥0.5). Since pseudotime was rank-transformed, its midpoint is 0.5.

Within each subtype, raw gene-expression counts were aggregated by donor into pseudobulk samples. Donor-level pseudobulk counts were normalized using TMM^54^. Genes that have zero expression in >99.9% of cells in the B-cell single-cell dataset or zero donor-level expression in more than 50% of donors within the corresponding subtype were removed. Gene expression values were then log transformed (log(1 + *x*)).

Expression values for each gene were scaled to have mean 0 and unit variance, and then used to compute pseudobulk expression PCs. The first 10 expression PCs, age, sex, and the first 5 genotype PCs were regressed from the normalized, log-transformed gene expression. The resulting residuals were transformed using a rank-based inverse-normal transformation before FUSION weight training.

For the Perez reference, FUSION weights^2^ were trained using variants within 500 kb of the gene TSS. The candidate prediction models were lasso, top1, and enet, evaluated using the standard fivefold cross-validation procedure implemented in FUSION. For the OneK1K analysis, we used FUSION’s default gene filter of heritability P-value<0.1. Because the Perez reference contained relatively few donors, we turned off the heritability filtered by setting hsq_set=1, allowing all genes to enter Stage-2 association analysis regardless of their expression heritability.

Replication analysis for FUSION was performed using the same approach as for Fast-TWiST, i.e., for genes with FDR<0.05 in OneK1K, we compute the proportion with evidence of association in Perez et al.

### Fast-TWiST analysis of T-cell GEPs in OneK1K

Functional gene expression programs (GEPs) were quantified in OneK1K T cells using starCAT version 1.0.10 with the fixed TCAT.V1 reference^27^. Of the 3,412 genes represented in the TCAT reference, 3,405 were available in the OneK1K expression data. starCAT computed GEP usage, defined as the normalized loadings of cells on the GEPs in a factorization model, jointly for 52 TCAT reference components across 882,630 T cells from 981 donors, including 12 T-cell subtypes. It constrained the values to be nonnegative and normalized within each cell so that the contributions of all reference components summed to one.

We ranked the 52 TCAT GEPs by their mean normalized GEP usage. Among the higher-ranking GEPs, we excluded components primarily associated with cell-type identity already annotated in OneK1K or technical variation (Translation, Mito, Poor-Quality). Due to correlations among GEPs, for each remaining candidate, cells were ranked by its usage, and the change in mean usage between the highest and lowest 10% of cells was calculated for all GEPs. A candidate was retained if its increase was at least 1.2 times the largest increase among the other GEPs, excluding Cytotoxic. This ensures that the GEP of interest is less confounded by correlated GEPs. The five highest-ranked GEPs meeting this criterion were Cytotoxic, IEG, BCL2/FAM13A, Th1-Like, and Th2-Resting.

For each selected GEP, normalized usage values were rank-scaled across all T cells to approximately uniform distribution from 0 to 1. Each functional GEP was analyzed independently using compressed FLASSO, with age, sex, top 10 expression PCs, and top 10 genotype PCs as covariates. Log library size was included as an offset in the Poisson model. Effects of SNPs on gene expression were represented using cubic B-splines with internal knots at 0.25, 0.50, and 0.75 and boundary knots at 0 and 1. We selected top 15,000 genes that had the largest mean raw UMI counts. Cis-SNP were restricted to 500 kb on either side of the TSS. Selection of tuning parameter λ was performed using five-fold cross-validation over a grid of 20 values.

Stage-2 association analysis was performed for CD, IBD, MS, RA, and SLE for each of the five GEPs, yielding 25 disease–GEP analyses. AS and T1DM had the smallest effective sample size (Ncase*Ncontrol/(Ncase+ Ncontrol)) and hence were not included. Within each disease and GEP, genes with FDR<0.05 were considered significant. We define MHC gene as those whose TSS were located between 25 and 34 Mb on chromosome 6 in GRCh37. We investigated GEP-sharing for MHC and non-MHC genes separately (Figure 4).

### Pathway over-representation analysis

For each combination of disease, GEP, and Fast-TWiST test (global or dynamic), we conducted pathway over-representation analysis for the set of significant genes (FDR<0.05). The background gene set comprises all genes with valid Fast-TWiST P-values that could be mapped to a gene symbol. Pathways were obtained from Gene Ontology Biological Process (GO-BP) 2026,^55,56^ using gene-set libraries downloaded from Enrichr.^57^ After intersection with the corresponding background genes, gene sets containing between 10 and 500 genes were retained. Enrichment was tested using a one-sided upper-tail hypergeometric test. P-values were adjusted across all eligible GO-BP pathways using the Benjamini–Hochberg procedure, separately for each disease, GEP, test, and MHC inclusion status.

Whole-genome and non-MHC pathway analyses were performed separately. For the non-MHC analysis, extended-MHC genes were removed from Fast-TWiST significant gene sets and background genes before recalculating pathway sizes, overlap counts, enrichment P-values, and pathway-level FDR. The original gene-level Fast-TWiST FDR values were retained. Genes contributing to the TCAT reference GEPs were retained in the primary enrichment analysis.

For each heatmap, GO-BP terms with nominal P < 0.05 in at least one of the five GEPs were considered separately for each disease. To remove highly overlapped terms for visualization purposes, GO terms were grouped by reference gene-set overlap (Jaccard similarity ≥ 0.25), equivalent to single-linkage hierarchical clustering with a distance cutoff of 0.75. Within each group, the representative was selected by prioritizing terms with pathway-level FDR < 0.05 in more GEPs. Ties were resolved by the numbers of GEPs meeting both FDR < 0.10, then by the number with nominal P < 0.05, the minimum P-value, the maximum enrichment ratio. Up to eight representatives were selected per disease, and duplicate terms were merged across diseases. This yielded 26 terms in the Figure 4 and 24, 39 and 36 terms in Supplementary Figures S7, S8 and S9.

### Overlap with published state-dependent T cell eQTLs

The dynamic gene set comprised genes with an original dynamic-association FDR<0.05 in at least one of the 25 disease-GEP analyses. Repeated discoveries across diseases and GEP were collapsed to 1,073 unique genes, which was then used to assess overlap with cell-state-dependent eQTLs in independent T cell datasets.

In Nathan et al,^15^ canonical correlation analysis (CCA) was used to identify shared patterns between gene expression and cell-surface protein measurements. Each pattern was represented by a continuous variable, called a canonical variate, which assigned a score to each cell. We used the joint genotype-by-state interaction tests across the first seven canonical variates from Supplementary Tables 20 and 34 of Nathan et al. Storey’s q-values were recalculated from the reported P-values separately for all genes in each table using the R package qvalue.^58^ Genes with q < 0.05 were retained for overlap with our dynamic gene set.

For Soskic et al,^34^ we used the dynamic eQTL results in Supplementary Table 8 of this paper. A gene was classified as having activation-dependent eQTL evidence if at least one tested lead SNP-gene pair had an FDR<0.05 for the linear or quadratic genotype-by-pseudotime interaction model in either naïve or memory CD4^+^ T cells. The reported FDR values were retained; they were calculated separately for the two interaction models within each cell population. This yielded 2,204 unique genes across the two populations.

### PBMC single-cell multiome data

Paired single-cell gene-expression and chromatin-accessibility data were obtained from the publicly available 10x Genomics PBMC Multiome dataset generated from cryopreserved PBMCs from one healthy 25-year-old female donor following granulocyte depletion. The dataset was generated using the Chromium Single-Cell Multiome ATAC + Gene Expression platform and was also used as a PBMC reference dataset in the development of SCENT.^59^

The processed dataset contained RNA measurements for 36,601 genes and chromatin-accessibility measurements for 132,291 peaks from the same 11,457 nuclei. The T-cell subset contained 5,942 nuclei spanning 12 T-cell annotations, all of which were included in the downstream regulatory analyses. The same five GEPs used in the OneK1K analysis were quantified in these T cells using starCAT with the fixed TCAT.V1 reference. For each GEP, usage values were rank-transformed across the 5,942 T cells to a scale from 0 to 1. Accessibility was analyzed separately for each GEP.

### Linking Stage-1 predictor SNPs to chromatin-accessibility peaks

For every gene-GEP pair with a dynamic association in at least one disease, we extracted SNPs with at least one nonzero coefficient (referred to as “predictor SNP”) in the corresponding Stage 1 expression-prediction model. We expanded the set of SNPs from predictor SNP to also include their LD proxies, defined as those with r^2^>0.80 in the 1000 Genomes European sample (N=489) with the predictor SNPs. Proxy SNPs were required to lie within 500 kb of both the predictor SNP and the TSS of the gene in GRCh37.

Predictor and proxy positions from OneK1K Fast-TWiST models were converted from GRCh37 to GRCh38 using the hg19ToHg38 liftOver chain; only unique mappings to the same chromosome were retained. A peak was eligible when it overlapped a predictor SNP or a proxy SNP within 500 kb of the gene transcription start site in GRCh38. After removing peaks that are detected in less than 5% of the analyzed cells, 27,356 peaks were retained for downstream analysis.

### Association between chromatin accessibility and GEP usage

For each of the 27,356 peaks, we tested the associated between chromatin accessibility against each of the five GEPs. Raw ATAC fragment counts were modeled using Poisson regression as follows:

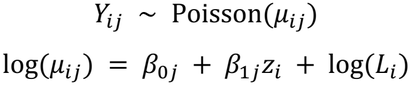

where *Y*_*ij*_ is the raw ATAC fragment count for peak *j* in cell *i, z*_*i*_ is the standardized, rank-transformed GEP usage, and *L*_*i*_ is the total raw ATAC fragment count for cell *i*.

P-value for the null hypothesis of *β*_*j*1_ = 0 was computed from a two-sided Wald test. Benjamini– Hochberg correction was applied jointly across all 136,780 peak–GEP tests. Peaks with a successful fit and FDR-adjusted P-value below 0.05 were classified as GEP-responsive. Both positive and negative accessibility associations qualified; no fold-change threshold was imposed.

A dynamic gene-GEP pair was considered to have ATAC support when at least one peak containing a predictor SNP or LD proxy was responsive to the same GEP. A gene was considered to have ATAC support if this criterion was met for any GEP.

### Visualization of GEP-responsive accessibility

For each locus, cells were divided into low, middle, and high groups based on their GEP ranks. Aggregated ATAC-seq tracks were generated using Signac,^60^ normalized by the total ATAC peak counts in each group, and smoothed using a 100-bp rolling sum.

### Example loci

To select example loci, we started with dynamically associated gene–GEP pairs and their nonzero Stage-1 predictor SNPs. We compared the predictors with cis-eQTL variants from Nathan or Soskic and genome-wide-significant variants from the corresponding disease GWAS (P < 5 × 10^−8^). The predictor, eQTL, and GWAS variants were required to be identical or in LD with one another (r^2^ ⩾ 0.6), calculated using 489 European individuals from the 1000 Genomes reference. We then examined nearby ATAC peaks and prioritized peaks containing SNPs linked to these variants and showing changes in accessibility across GEP ranks. ORMDL3-SLE and PTGER4-CD were selected for satisfying all the criteria above.

## Data Availability

OneK1K single-cell RNA-seq and genotype data are available through GEO GSE196830. The Perez et al. single-cell RNA-seq data are available through GEO GSE174188, and the genotype data are available through dbGaP phs002812.v1.p1. Published dynamic eQTL results were obtained from Supplementary Tables 20 and 34 (https://media.springernature.com/original/springer-static/esm/art%3A10.1038%2Fs41586-022-04713-1/MediaObjects/41586_2022_4713_MOESM3_ESM.xlsx) of Nathan et al. (2022) and Supplementary Table 8 (https://static-content.springer.com/esm/art%3A10.1038%2Fs41588-022-01066-3/MediaObjects/41588_2022_1066_MOESM4_ESM.xlsx) of Soskic et al. (2022).

For locus-level comparisons, eQTL summary statistics were obtained from the eQTL Catalogue for Nathan et al. (https://ftp.ebi.ac.uk/pub/databases/spot/eQTL/sumstats/QTS000040/) and from Zenodo for Soskic et al. (https://zenodo.org/records/6006796). The PBMC multiome dataset is available from 10x Genomics PBMC (https://www.10xgenomics.com/datasets/pbmc-from-a-healthy-donor-granulocytes-removed-through-cell-sorting-10-k-1-standard-2-0-0).

Publicly available European-ancestry Million Veteran Program summary statistics were obtained from dbGaP accession phs002453.v1.p1 MVP summary-results study (https://www.ncbi.nlm.nih.gov/projects/gap/cgi-bin/study.cgi?study_id=phs002453.v1.p1) for ankylosing spondylitis (Phe_715_2), inflammatory bowel disease (Phe_555), multiple sclerosis (Phe_335), and rheumatoid arthritis (Phe_714_1). FinnGen Release 12 summary statistics (Kurki et al., 2023) were used for Crohn’s disease, type 1 diabetes, and systemic lupus erythematosus FinnGen Release 12 data access (https://finngen.gitbook.io/documentation/data-download): Crohn’s disease K11_CD_STRICT2 summary statistics, type 1 diabetes T1D summary statistics, and systemic lupus erythematosus SLE_FG summary statistics.

## Code Availability

Fast-TWiST has been added to the TWiST repository on GitHub: https://github.com/gqi/TWiST.

## Acknowledgements

J.C. and G.Q. were supported by NIH/NHGRI award K01HG013983.

