## Supplementary Figures and Tables for "Scalable cell-state-resolved TWAS identified dynamic autoimmune disease genes along functional programs"

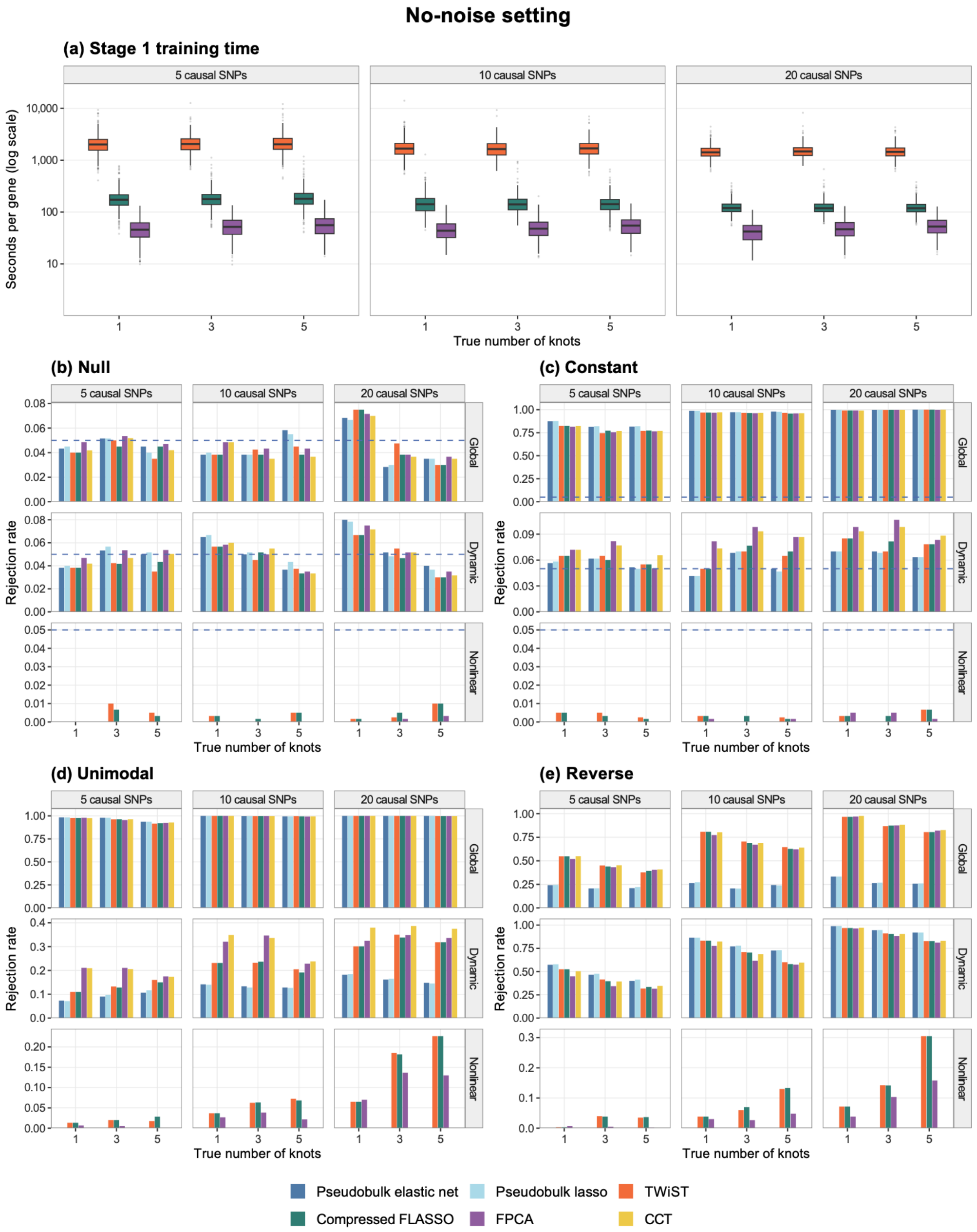


**Supplementary Figure S1. Computation time, type I error, and power in simulations without added noise.** (a) Stage-1 training time per gene for TWiST, compressed FLASSO, and FPCA. (b) Type I error for global, dynamic, and nonlinear tests in null simulations. (c) Power of the global test and type I error of the dynamic and nonlinear tests in the constant scenario. (d–e) Power in the unimodal and reverse scenarios. Six methods were compared: pseudobulk elastic net, pseudobulk lasso, TWiST, compressed FLASSO, FPCA, and the Cauchy combination test (CCT) of compressed FLASSO and FPCA. Number of causal SNPs in the cis-region of the gene varies among 5, 10, and 20. True SNP effects on expression and true gene effect on trait $\beta(t)$ are B-spline functions between 0 and 1 with varying number of knots: 1, 3, and 5. All other settings are as in Figure 2. Blue dashed lines represent significant threshold p = 0.05.

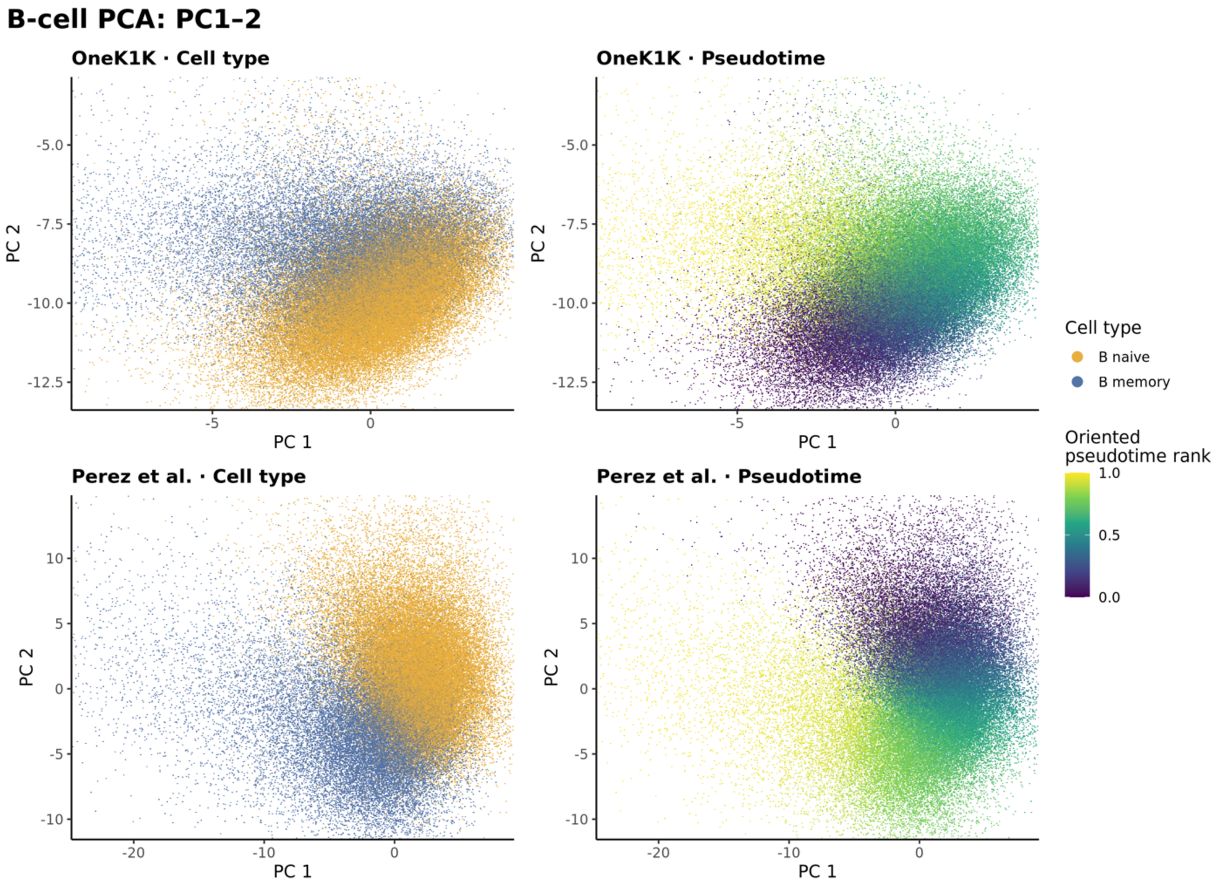


**Supplementary Figure S2. Naïve to Memory B cell PCA in OneK1K and Perez.** Left panels show Onek1k and Perez B-naive/B-memory annotations; right panels show Slingshot pseudotime oriented from naive to memory. The pseudotime is scaled from 0 (early) to 1 (late). Each cohort is shown in its own PCA space.

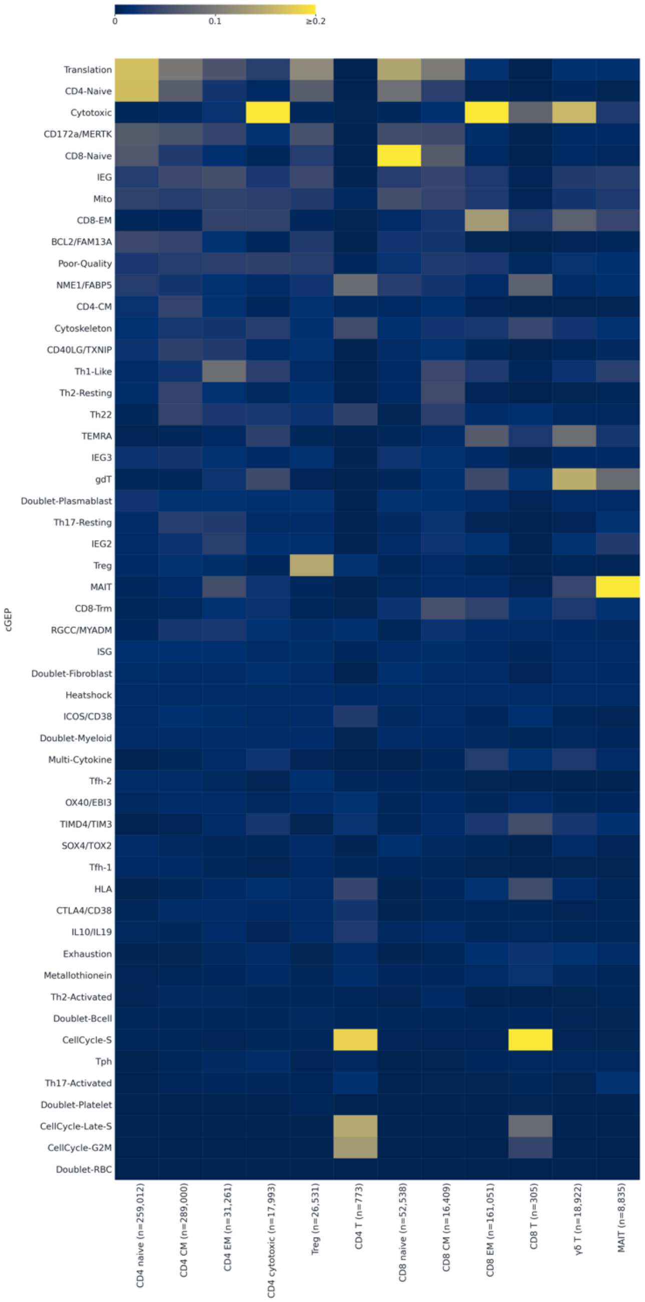


**Supplementary Figure S3. Mean GEP usage across OneK1K T-cell types.** The heatmap shows the mean usage of 52 TCAT gene expression programs (GEPs), compted using starCAT, across 12 Onek1k annotated T-cell types. Programs are ordered from top to bottom by decreasing mean usage across all T cells. Colors show mean usage on a linear scale from 0 to 0.2, with values ≥0.2 shown at the maximum color intensity.

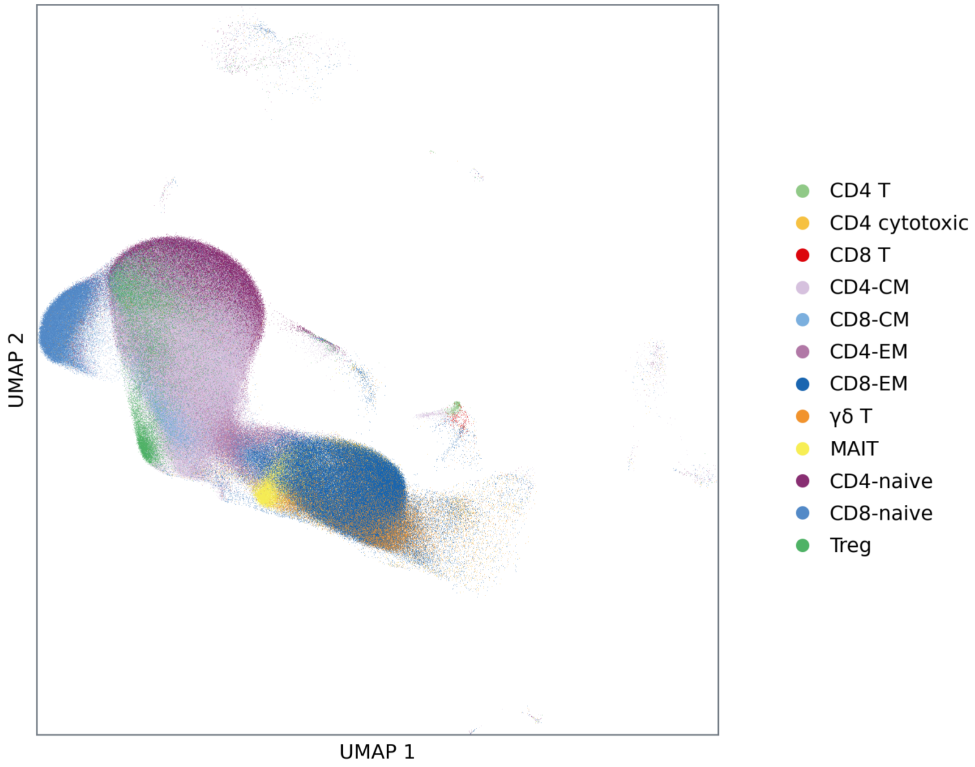


**Supplementary Figure S4. UMAP of OneK1K T cells colored by subtype.** Abbreviations: CM, central memory; EM, effector memory; MAIT, mucosal-associated invariant T cell; Treg, regulatory T cell.

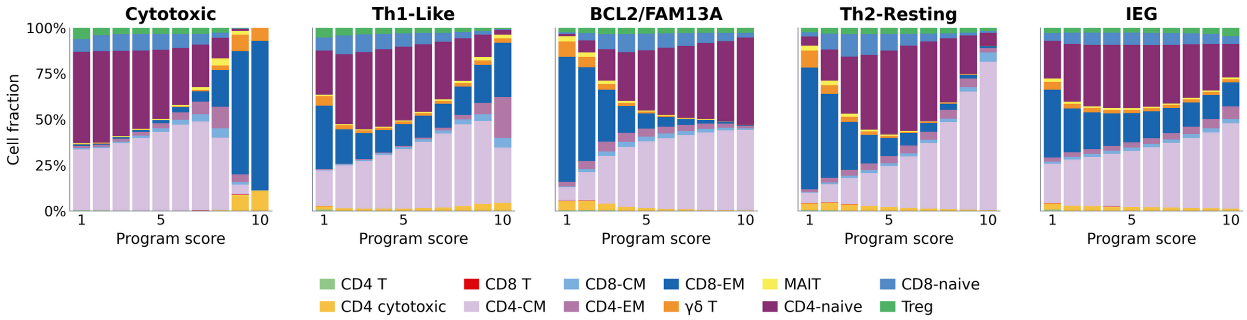


**Supplementary Figure S5. T-cell subtype composition across gene expression program (GEP) usage values.** For each of the five GEPs, cells were ranked by their usage values and divided into ten bins. Colors correspond to those in the UMAP in Supplementary Figure S4.

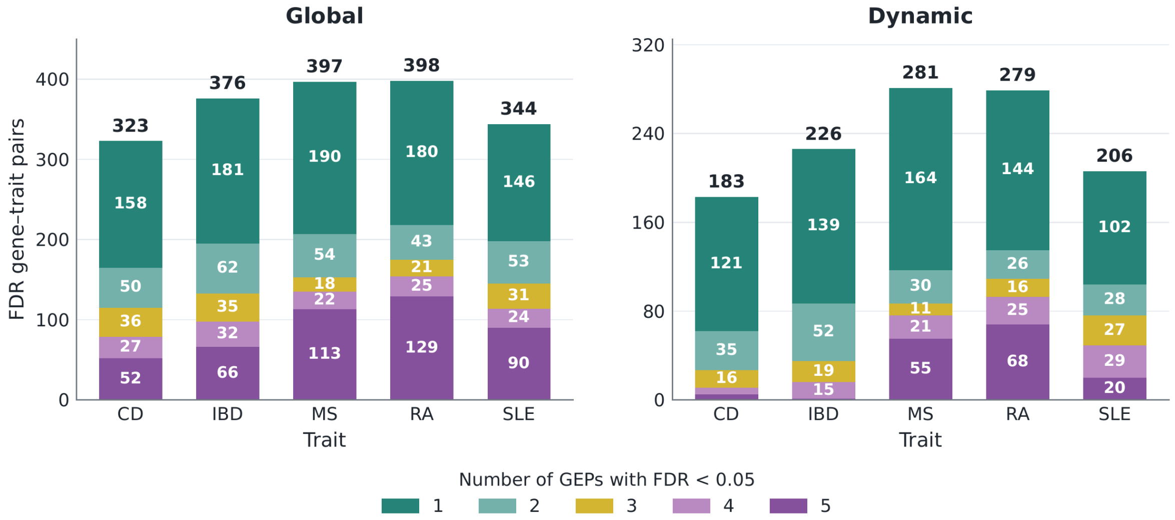


**Supplementary Figure S6. Number of significant (FDR < 0.05) genes associated with each disease by GEP sharing level with both MHC and non-MHC genes are included.** Colors indicate genes shared by number of gene expression programs.

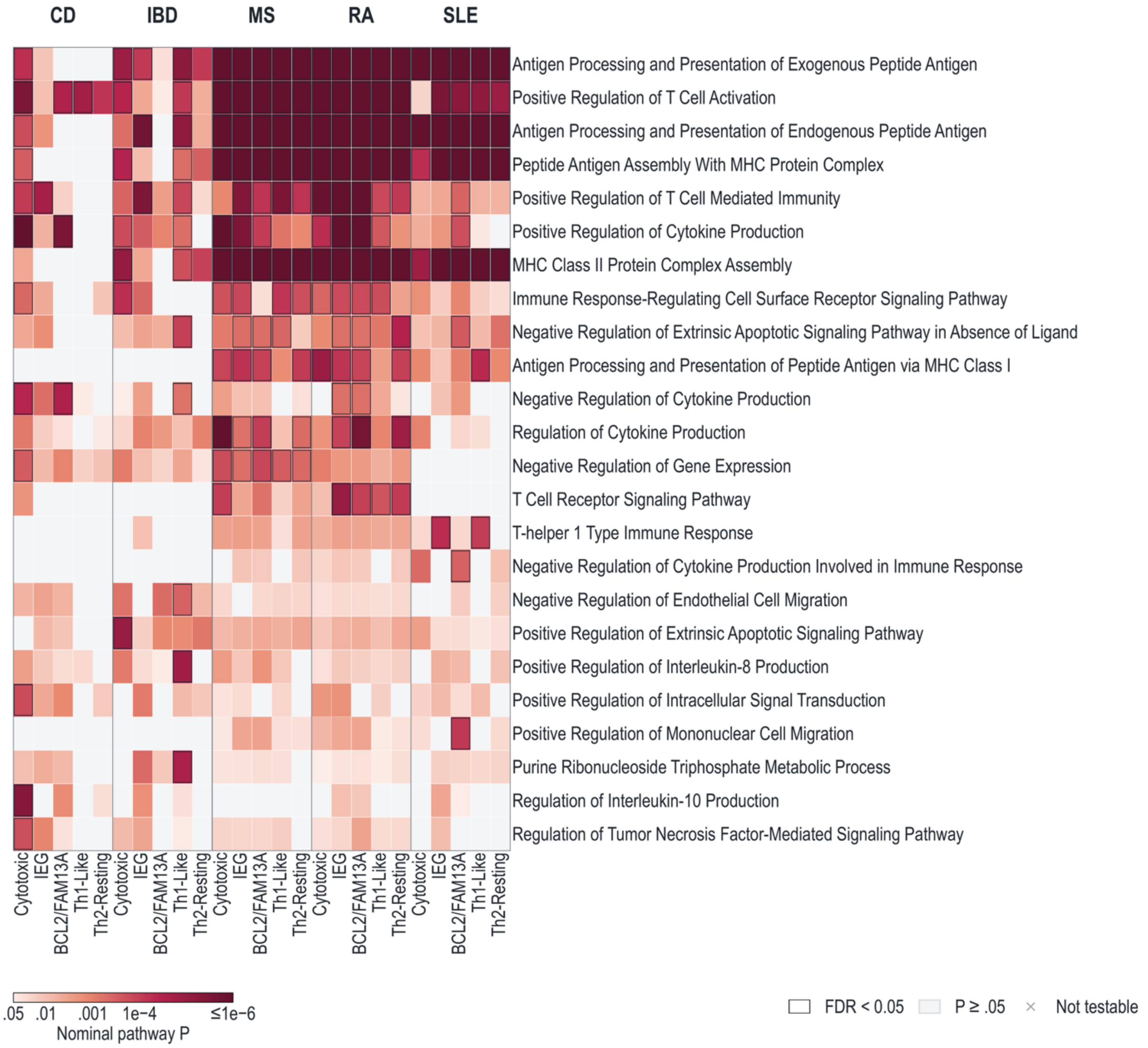


**Supplementary Figure S7. Over-representation analysis (ORA) of dynamic genes, including MHC.** ORA was performed using genes with significant dynamic associations (FDR < 0.05) in each of the 25 disease–GEP combinations. For each disease, GO Biological Process terms with nominal P < 0.05 in at least one GEP were grouped by reference gene-set overlap (Jaccard similarity ≥ 0.25). One representative was selected per group, prioritizing terms with pathway-level FDR < 0.05 in more GEPs. Up to eight representatives per disease were combined, yielding 24 unique terms. Rows are ordered by the number of diseases with pathway FDR < 0.05 in at least one GEP, then by the number of significant disease–GEP combinations. Ties are ordered alphabetically. Colors indicate nominal hypergeometric P values, with P ≤ $10^{-6}$ shown at the maximum color intensity. Gray indicates P ≥ 0.05, and black outlines indicate Benjamini–Hochberg-adjusted P < 0.05. A cross (×) indicates a pathway that was not evaluated because its size in the corresponding tested-gene background was outside the 10–500-gene range.

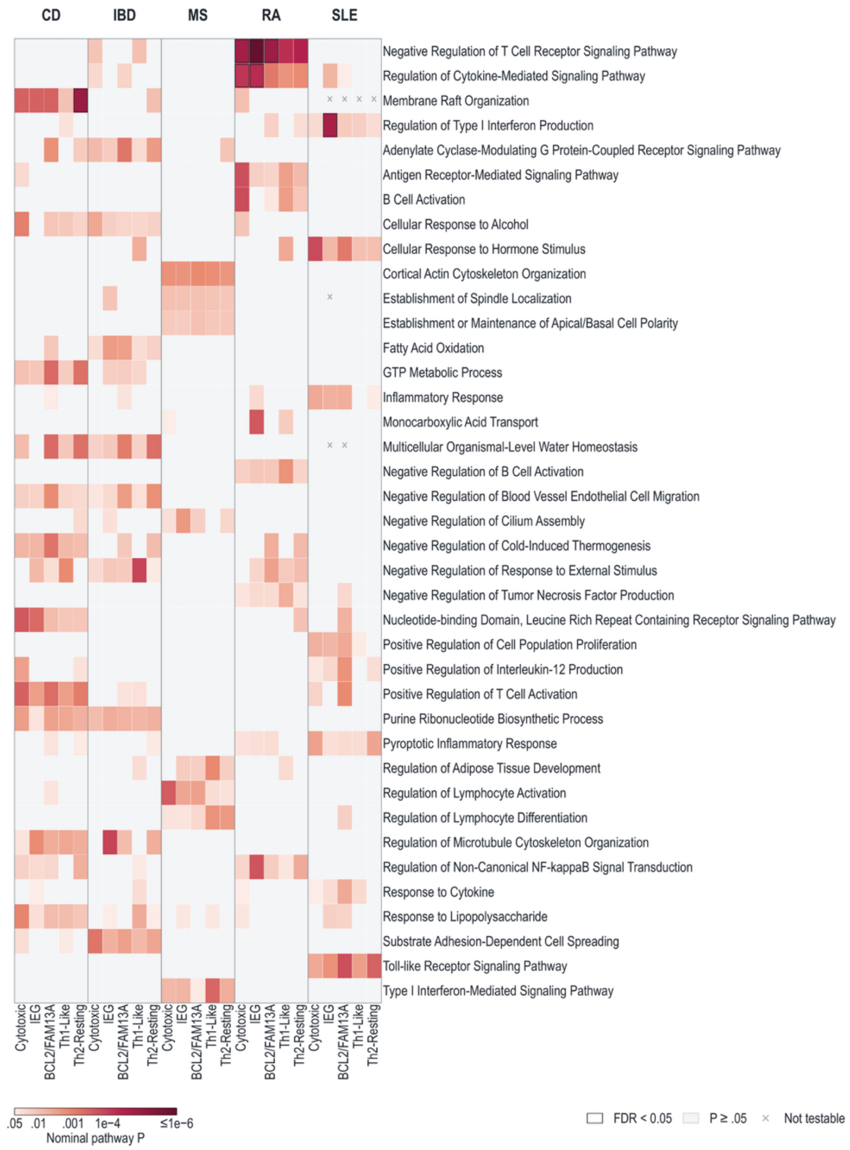


**Supplementary Figure S8. Over-representation analysis (ORA) of globally significant genes, excluding MHC.** ORA was performed using genes with significant global associations (FDR < 0.05) in each of the 25 disease–GEP combinations. Genes in the extended MHC region were removed from both the associated-gene set and the corresponding tested-gene background before ORA. For each disease, GO Biological Process terms with nominal P < 0.05 in at least one GEP were grouped by reference gene-set overlap (Jaccard similarity ≥ 0.25). Rows are ordered by the number of diseases with pathway FDR < 0.05 in at least one GEP, then by the number of significant disease–GEP combinations. Ties are ordered alphabetically. Up to eight representatives per disease were combined, yielding 39 unique terms. Colors indicate nominal hypergeometric P values, with $P \leq 10^{-6}$ shown at the maximum color intensity. Gray indicates P ≥ 0.05, and black outlines indicate Benjamini–Hochberg-adjusted P < 0.05. A cross (×) indicates that fewer than 10 pathway genes were present in the corresponding background after MHC removal.

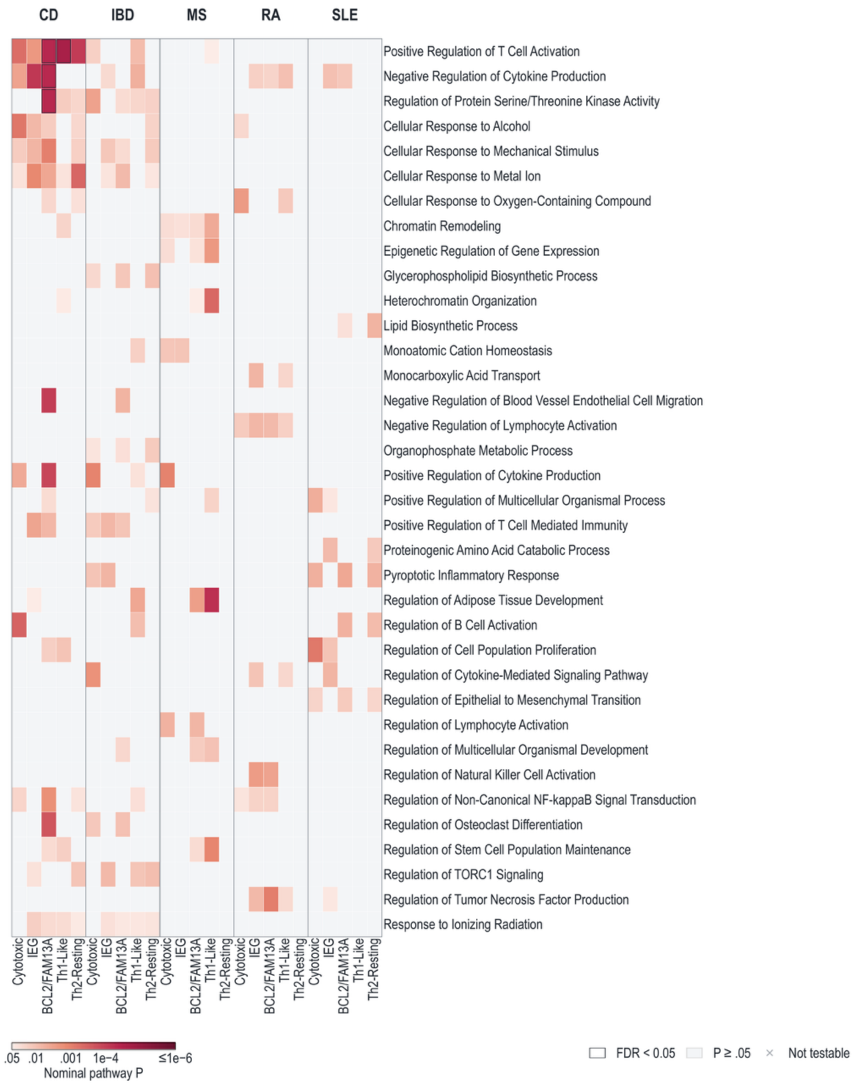


**Supplementary Figure S9. Over-representation analysis (ORA) of dynamic genes, excluding MHC.** ORA was performed using genes with significant dynamic associations (FDR < 0.05) in each of the 25 disease–GEP combinations. Genes in the extended MHC region were removed from both the associated-gene set and the corresponding tested-gene background before ORA. For each disease, GO Biological Process terms with nominal P < 0.05 in at least one GEP were grouped by reference gene-set overlap (Jaccard similarity ≥ 0.25). One representative was selected per group, prioritizing terms with pathway-level FDR < 0.05 in more GEPs. Up to eight representatives per disease were combined, yielding 36 unique terms. Rows are ordered by the number of diseases with pathway FDR < 0.05 in at least one GEP, then by the number of significant disease–GEP combinations. Ties are ordered alphabetically. Colors indicate nominal hypergeometric P values, with P ≤ $10^{-6}$ shown at the maximum color intensity. Gray indicates P ≥ 0.05, and black outlines indicate Benjamini–Hochberg-adjusted P < 0.05. A cross (×) indicates a pathway that was not evaluated because its size in the corresponding tested-gene background was outside the 10–500-gene range.

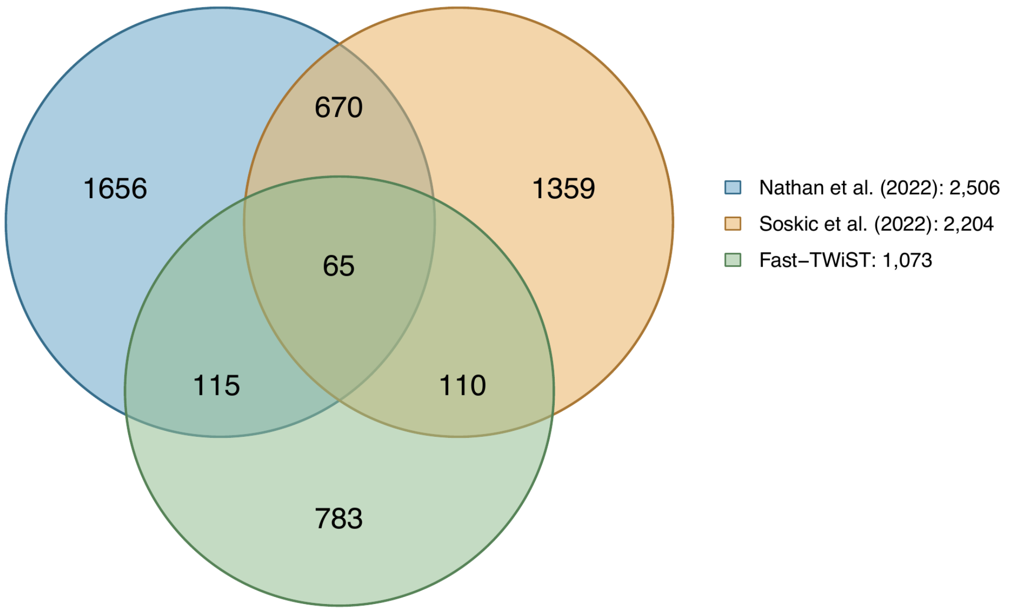


**Supplementary Figure S10. Overlap of Fast-TWiST dynamic genes with published state-dependent eQTL genes.** The Venn diagram shows the overlap between genes with a significant dynamic Fast-TWiST association in at least one disease-GEP analysis (FDR < 0.05) and genes with state-dependent eQTLs in Nathan et al. (2022; recalculated Storey q < 0.05) and Soskic et al. (2022; reported FDR < 0.05). Numbers represent unique genes in each region.

| Disease/ Program | Cytotoxic | IEG | BCL2/FAM13A | Th1-Like | Th2-Resting | Total |
|---|---|---|---|---|---|---|
| CD | 175 / 80 | 178 / 57 | 177 / 75 | 197 / 73 | 193 / 86 | 920 / 371 |
| IBD | 255 / 117 | 212 / 84 | 222 / 100 | 239 / 96 | 198 / 85 | 1,126 / 482 |
| MS | 272 / 160 | 264 / 156 | 248 / 155 | 263 / 150 | 253 / 151 | 1,300 / 772 |
| RA | 274 / 194 | 250 / 160 | 247 / 161 | 302 / 185 | 276 / 159 | 1,349 / 859 |
| SLE | 247 / 124 | 216 / 110 | 212 / 116 | 216 / 128 | 240 / 130 | 1,131 / 608 |
| Total | 1,223 / 675 | 1,120 / 567 | 1,106 / 607 | 1,217 / 632 | 1,160 / 611 | 5,826 / 3,092 |

**Supplementary Table S1. Global and dynamic associations across diseases and T-cell gene expression programs.** Each entry shows the numbers of significant genes from the global and dynamic tests, respectively (global / dynamic). FDR was controlled using the Benjamini–Hochberg procedure separately for each disease–program pair and test (FDR < 0.05).
