## Supplementary Methods for "Scalable cell-state-resolved TWAS identified dynamic autoimmune disease genes along functional programs"

#### S1. Compressed FLASSO: fitting the group elastic-net model using individual-level summaries

A direct fit of the group elastic-net model in the main Methods, restated in (S1) below, multiplies each SNP's cell-level design block by the current working residuals, so its cost grows with the number of cells. All cells from the same individual share the same genotype, and compressed FLASSO uses this structure to carry out the same iterations using summaries computed at the individual level. This section derives the coordinate-descent iterations of Algorithm 1, shows which individual-level summaries they require, and explains how each step of the algorithm follows.

There are $N=\sum_{i=1}^{I} J_i$ cells from $I$ individuals. $\boldsymbol{\phi}(t)=\left(\phi_1(t),\dots,\phi_M(t)\right)^{\top}$ is the Stage 1 B-spline basis, and $\mathbf{G}_k$ is the $N\times M$ design block for SNP $k$, whose row for cell $j$ of individual $i$ is $g_{ik}\boldsymbol{\phi}\left(t_{ij}\right)^{\top}$. The transformed block is $\widetilde{\mathbf{G}}_k=(\mathbf{G}_k-\mathbf{1}\mathbf{u}_k^{\top})\mathbf{T}_k$ with $\widetilde{\mathbf{G}}_k^{\top}\widetilde{\mathbf{G}}_k=N\mathbf{I}_M$ (the centering vector $\mathbf{u}_k$ and the transformation $\mathbf{T}_k$ are defined below), $\widetilde{\mathbf{w}}_k$ is the coefficient vector in the transformed block, and $\mathbf{w}_k=\mathbf{T}_k\widetilde{\mathbf{w}}_k$ is the coefficient vector in the original spline basis. We write $\mathbf{x}=\left(x_{ij}\right)$ for the vector of expression counts, $\boldsymbol{\eta}=\left(\eta_{ij}\right)$ for the vector of linear predictors, $\eta_{ij}=\hat{o}_{ij}+c+\sum_{k=1}^{K}[\widetilde{\mathbf{G}}_k\widetilde{\mathbf{w}}_k]_{ij}$, and $\boldsymbol{\mu}=\exp(\boldsymbol{\eta})$ for the fitted means, where $\hat{o}_{ij}$ is the fixed offset from the covariate-only model in the main Methods.

*Derivatives and coordinate updates.*

As in the main Methods, compressed FLASSO minimizes the average negative Poisson log-likelihood plus the group elastic-net penalty,

$$\min_{c,\{\widetilde{\mathbf{w}}_k\}_{k=1}^{K}}\left\{-\frac{1}{N}\ell(c,\widetilde{\mathbf{w}}_1,\dots,\widetilde{\mathbf{w}}_K\mid\mathbf{x},\hat{\mathbf{o}})+\lambda\sum_{k=1}^{K}\sqrt{M}\left[\rho\parallel\widetilde{\mathbf{w}}_k\parallel_2+\frac{1-\rho}{2}\parallel\widetilde{\mathbf{w}}_k\parallel_2^2\right]\right\},\qquad(\mathrm{S1})$$

where the Poisson log-likelihood is $\ell=\sum_{i,j}\left(x_{ij}\eta_{ij}-\mu_{ij}\right)$, For the uncentered blocks $\mathbf{G}_k$ (centering only shifts the intercept), the derivatives with respect to the intercept and the original spline coefficients $\mathbf{w}_k$ are

$$\frac{\partial\ell}{\partial c}=\sum\nolimits_{i,j}\left(x_{ij}-\mu_{ij}\right),\qquad\frac{\partial\ell}{\partial\mathbf{w}_k}=\mathbf{G}_k^{\top}(\mathbf{x}-\boldsymbol{\mu})=\sum\nolimits_{i}g_{ik}\sum\nolimits_{j}\boldsymbol{\phi}\left(t_{ij}\right)\left(x_{ij}-\mu_{ij}\right),$$

$$\frac{\partial^2\ell}{\partial\mathbf{w}_k\,\partial\mathbf{w}_k^{\top}}=-\mathbf{G}_k^{\top}\mathrm{diag}(\boldsymbol{\mu})\mathbf{G}_k=-\sum\nolimits_{i}g_{ik}^2\sum\nolimits_{j}\mu_{ij}\;\boldsymbol{\phi}\left(t_{ij}\right)\boldsymbol{\phi}\left(t_{ij}\right)^{\top}.$$

Following grpreg[1], the curvature $\mu_{ij}$ in the second derivative is replaced by its upper bound $a=\max_{i,j}\mu_{ij}$, which gives $-a\sum_i g_{ik}^2\,\mathbf{M}_i$ with $\mathbf{M}_i=\sum_j\boldsymbol{\phi}\left(t_{ij}\right)\boldsymbol{\phi}\left(t_{ij}\right)^{\top}$; this is why $\mathbf{M}_i$ is precomputed. In the transformed block the same quantities are $\partial\ell/\partial\widetilde{\mathbf{w}}_k=\widetilde{\mathbf{G}}_k^{\top}(\mathbf{x}-\boldsymbol{\mu})$ and $-\widetilde{\mathbf{G}}_k^{\top}\mathrm{diag}(\boldsymbol{\mu})\widetilde{\mathbf{G}}_k$, and the bound becomes $-a\,\widetilde{\mathbf{G}}_k^{\top}\widetilde{\mathbf{G}}_k=-aN\mathbf{I}_M$. Define the working residuals $r_{ij}=\left(x_{ij}-\mu_{ij}\right)/a$ and $\mathbf{r}=\left(r_{ij}\right)$. The coordinate update of grpreg for $\widetilde{\mathbf{w}}_k$, with the other coefficients held fixed, is

$$\mathbf{z}_k = \widetilde{\mathbf{w}}_k + \frac{1}{N}\widetilde{\mathbf{G}}_k^{\top}\mathbf{r}, \qquad \widetilde{\mathbf{w}}_k \leftarrow \frac{\left(a \parallel \mathbf{z}_k \parallel_2 - \lambda\sqrt{M}\rho\right)_+}{a\left(1 + \lambda\sqrt{M}(1-\rho)\right)} \frac{\mathbf{z}_k}{\parallel \mathbf{z}_k \parallel_2}, \tag{S2}$$

where $(u)_+ = \max(u, 0)$, $\lambda$ and $\rho$ are the penalty parameters in **Error! Reference source not found.**, and the update of the unpenalized intercept is $c \leftarrow c + N^{-1}\sum_{i,j} r_{ij}$. One outer iteration recalculates $\boldsymbol{\mu}$, $a$ and $\mathbf{r}$ at the current coefficients (lines 4–5 of Algorithm 1) and then performs one inner cycle of these updates over the intercept and $k = 1, \dots, K$. Within the cycle $a$ is held fixed because it is the curvature of the quadratic approximation constructed at the start of the outer iteration, and $\mathbf{r}$ is updated after each coefficient change (Algorithm 1). Outer iterations are repeated until the largest coefficient change within an iteration is below $10^{-4}$.

The quantities required are therefore the column means and cross-products of $\mathbf{G}_k$, which determine $\mathbf{u}_k$, $\mathbf{T}_k$, and $\widetilde{\mathbf{G}}_k^{\top}\mathbf{r}$ at each update; both are obtained from individual-level summaries as follows.

*Basis summaries and coefficient transformation.*

Before optimization, we calculate

$$\mathbf{S}_i = \sum_{j=1}^{J_i} \boldsymbol{\phi}(t_{ij}), \qquad \mathbf{M}_i = \sum_{j=1}^{J_i} \boldsymbol{\phi}(t_{ij})\boldsymbol{\phi}(t_{ij})^{\top}, \qquad i = 1, \dots, I.$$

These quantities depend on the cells and basis functions, but not on genotype or fitted expression. They are calculated once for each gene. For SNP $k$, the vector of column means is $\mathbf{u}_k = N^{-1}\sum_i g_{ik}\,\mathbf{S}_i$, and the matrix of centered column cross-products is

$$\boldsymbol{\Gamma}_k = \sum_{i=1}^{I} g_{ik}^2\,\mathbf{M}_i - N\mathbf{u}_k\mathbf{u}_k^{\top}.$$

Here $\sum_i g_{ik}^2\,\mathbf{M}_i = \mathbf{G}_k^{\top}\mathbf{G}_k$, and $\boldsymbol{\Gamma}_k$ is the cross-product matrix of the centered block $\mathbf{G}_k - \mathbf{1}\mathbf{u}_k^{\top}$. This small $M \times M$ matrix provides the information needed to scale and orthogonalize the SNP's design block. Let $\mathbf{T}_k$ denote the resulting transformation: from the eigendecomposition $\boldsymbol{\Gamma}_k = \mathbf{V}_k\mathrm{diag}(d_{k1}, \dots, d_{kM})\mathbf{V}_k^{\top}$, $\mathbf{T}_k = \mathbf{V}_k\mathrm{diag}\left(\sqrt{N/d_{k1}}, \dots, \sqrt{N/d_{kM}}\right)$, so that $\widetilde{\mathbf{G}}_k = (\mathbf{G}_k - \mathbf{1}\mathbf{u}_k^{\top})\mathbf{T}_k$ satisfies $\widetilde{\mathbf{G}}_k^{\top}\widetilde{\mathbf{G}}_k = N\mathbf{I}_M$, where $\mathbf{1}$ is a vector of $N$ ones. After fitting, the original spline coefficients are recovered as $\mathbf{w}_k = \mathbf{T}_k\widetilde{\mathbf{w}}_k$. Centering changes only the intercept contribution and does not change the recovered SNP-effect curve.

*Individual-level computation of the updates.*

A single pass over the cells gives $\mathbf{H}_i = \sum_j \boldsymbol{\phi}(t_{ij})r_{ij}$ for each individual. The residual cross-product for the transformed block of SNP $k$ in the update for $\mathbf{z}_k$ is then

$$\widetilde{\mathbf{G}}_k^{\top}\mathbf{r} = \mathbf{T}_k^{\top}\left\{\sum_{i=1}^{I} g_{ik}\,\mathbf{H}_i - \mathbf{u}_k\sum_{i,j} r_{ij}\right\}.$$

In grpreg, $\widetilde{\mathbf{G}}_k^\top \mathbf{r}$ is computed as a sum over all $N$ cells at every SNP update. Here it is a sum over $I$ individuals, and the $\mathbf{H}_i$ are computed in a single pass over the cells and shared by all SNPs. After the intercept update, $\sum_{i,j} r_{ij} = 0$, and it remains zero after every SNP update because the transformed blocks are centered ($\sum_{i,j} \Delta\eta_{ij} = 0$ below), so the second term vanishes throughout the cycle. Following the update of $\widetilde{\mathbf{w}}_k$, let $\Delta\mathbf{w}_k = \mathbf{T}_k \Delta\widetilde{\mathbf{w}}_k$ be the change in the original coefficients. The linear predictor of each cell changes by $\Delta\eta_{ij} = g_{ik}\boldsymbol{\phi}(t_{ij})^\top \Delta\mathbf{w}_k - \mathbf{u}_k^\top \Delta\mathbf{w}_k$, and since $a$ is fixed, $r_{ij} \leftarrow r_{ij} - \Delta\eta_{ij}$. Summing over the cells of individual $i$ gives

$$\mathbf{H}_i \leftarrow \mathbf{H}_i - g_{ik}\mathbf{M}_i \Delta\mathbf{w}_k + \mathbf{S}_i(\mathbf{u}_k^\top \Delta\mathbf{w}_k).$$

The last term accounts for centering. The solver also maintains the individual-level predictor coefficients $\mathbf{F}_i = \sum_k g_{ik}\,\mathbf{w}_k$, along with the intercept and total working residual, so that $\eta_{ij} = \hat{o}_{ij} + c - \sum_k \mathbf{u}_k^\top \mathbf{w}_k + \boldsymbol{\phi}(t_{ij})^\top \mathbf{F}_i$ is obtained in one pass over the cells. These quantities allow the fitted values and residual summaries to be updated without revisiting every cell for each SNP. Cell-level means and working residuals are recalculated at the next outer iteration. The static summaries $\mathbf{S}_i$ and $\mathbf{M}_i$ therefore do not replace the expression counts or the changing residual summaries.

Algorithm 1 summarizes the resulting procedure.

**Algorithm 1. Compressed FLASSO for a fixed penalty $\lambda$.**

---

Precompute $\mathbf{S}_i$, $\mathbf{M}_i$ for each individual, and $\mathbf{u}_k$, $\mathbf{T}_k$ for each SNP<br>
Initialize $c \leftarrow 0$ and $\widetilde{\mathbf{w}}_k \leftarrow \mathbf{0}$<br>
**repeat**<br>
    $\mu_{ij} \leftarrow \exp\left(\hat{o}_{ij} + c + \sum_k [\widetilde{\mathbf{G}}_k \widetilde{\mathbf{w}}_k]_{ij}\right)$ for every cell *(one pass over cells)*<br>
    $a \leftarrow \max_{i,j}\mu_{ij}$; $r_{ij} \leftarrow (x_{ij} - \mu_{ij})/a$ *(curvature and working residuals)*<br>
    $\mathbf{H}_i \leftarrow \sum_j \boldsymbol{\phi}(t_{ij})\, r_{ij}$ for each individual<br>
    $\delta \leftarrow N^{-1}\sum_{i,j} r_{ij}$; $c \leftarrow c + \delta$; $\mathbf{H}_i \leftarrow \mathbf{H}_i - \delta\,\mathbf{S}_i$ for each individual *(intercept)*<br>
    **for** $k = 1, \ldots, K$ **do** *(individual-level updates only)*<br>
        $\mathbf{z}_k \leftarrow \widetilde{\mathbf{w}}_k + \frac{1}{N}\,\mathbf{T}_k^\top \sum_i g_{ik}\,\mathbf{H}_i$ *(equals $\widetilde{\mathbf{w}}_k + \widetilde{\mathbf{G}}_k^\top \mathbf{r}/N$ because $\sum_{i,j} r_{ij} = 0$)*<br>
        $\widetilde{\mathbf{w}}_k^{\mathrm{new}} \leftarrow \frac{(a\|\mathbf{z}_k\|_2 - \lambda\sqrt{M}\rho)_+}{a\left(1+\lambda\sqrt{M}(1-\rho)\right)} \cdot \frac{\mathbf{z}_k}{\|\mathbf{z}_k\|_2}$ *(group soft-threshold; $(u)_+ = \max(u, 0)$)*<br>
        $\Delta\mathbf{w}_k \leftarrow \mathbf{T}_k(\widetilde{\mathbf{w}}_k^{\mathrm{new}} - \widetilde{\mathbf{w}}_k)$; $\widetilde{\mathbf{w}}_k \leftarrow \widetilde{\mathbf{w}}_k^{\mathrm{new}}$<br>
        $\mathbf{H}_i \leftarrow \mathbf{H}_i - g_{ik}\mathbf{M}_i\Delta\mathbf{w}_k + \mathbf{S}_i(\mathbf{u}_k^\top \Delta\mathbf{w}_k)$ for each individual<br>
    **end for**<br>
**until** the largest change in $c$ and $\widetilde{\mathbf{w}}_k$ within an iteration is below $10^{-4}$<br>
**return** $\mathbf{w}_k = \mathbf{T}_k\widetilde{\mathbf{w}}_k$ for $k = 1, \ldots, K$

---

*Computational cost.*

For a direct fit, constructing the $N \times KM$ design matrix requires $O(NKM)$ operations, and computing the $K$ cross-product matrices $\mathbf{G}_k^\top \mathbf{G}_k$, each $M \times M$ with entries summed over $N$ cells, requires $O(NKM^2)$ operations. Compressed FLASSO instead computes the individual-level basis summaries $\mathbf{M}_i$ for all individuals in $O(NM^2)$ operations, forms cross-product matrices $\sum_i g_{ik}^2 \mathbf{M}_i$ for $K$ SNPs in $O(KIM^2)$ operations and obtains $\mathbf{T}_k$ from the eigendecomposition of each $\mathbf{\Gamma}_k$ in $O(KM^3)$ operations. During fitting, refreshing the cell-level quantities requires $O(NM)$ operations per outer iteration (lines 4-7 of Algorithm 1), while a SNP update and the associated summary updates require $O(IM^2)$ operations (lines 9-12). In comparison, a direct fit computes $\widetilde{\mathbf{G}}_k^\top \mathbf{r}$ by summing over $N$ cells, which costs $O(NM)$ per SNP update. Because $IM^2 < NM$ whenever $M$ is smaller than the average number of cells per individual, $N/I$, the main reduction is therefore in the repeated SNP-specific calculations, which depend on the number of individuals rather than the number of cells.

*Numerical validation.*

We validated the implementation against two independent reference solvers: a cell-level solver that reconstructs each SNP's design block on demand, and a solver operating on an explicitly constructed, pre-standardized and pre-orthogonalized design matrix that mirrors the reference grpreg implementation (family = "poisson", penalty = "grLasso", alpha = $\rho$). Fitted coefficients agreed with both reference solvers to numerical precision on simulated data. On OneK1K B cells, computing $\mathbf{T}_k$ from the eigendecomposition of $\mathbf{\Gamma}_k$ and from the singular value decomposition of the explicitly constructed centered design block gave the same fits.

### S2. Likelihood based on GWAS summary statistics

We follow the TWiST derivation[2], allowing the Stage 1 basis to be either the B-spline basis of TWiST and compressed FLASSO or the eigenfunctions estimated by FPCA. For this derivation, we denote either basis by $\{\phi_m(t)\}_{m=1}^M$; for FPCA, $M = L$ and $\phi_m(t) = \varphi_m(t)$ in the main Methods. We use the following notation:

- $y_a$: the trait of individual $a$ in the GWAS sample, after adjustment for covariates and standardization to mean zero and variance one.
- $g_{ak}$: the genotype of individual $a$ at cis-SNP $k$, standardized to mean zero and variance one. The GWAS individuals need not be the individuals used for Stage 1 training.
- $w_{km}$: the fitted Stage 1 coefficient for SNP $k$ and basis function $m$. Hats are omitted from the fitted coefficients in this derivation for readability.
- $v_a(t) = \sum_{k=1}^K \sum_{m=1}^M w_{km}\, \phi_m(t) g_{ak}$: the GReX trajectory obtained from the fitted Stage 1 model.
- $\beta(t) = \beta_{l0} + \beta_{l1} t + \sum_{m'=1}^{M'} \beta_{m'} \psi_{m'}(t)$: the effect of GReX on the trait, with the Stage 2 B-spline functions $\psi_{m'}(t)$ defined in the main Methods.

Let $W$ be the $K \times M$ matrix with entries $w_{km}$, and define the column vectors $\boldsymbol{\beta}_l = (\beta_{l0}, \beta_{l1})^\top$, $\boldsymbol{\beta} = (\beta_1, \dots, \beta_{M'})^\top$, and $\mathbf{g}_a = (g_{a1}, \dots, g_{aK})^\top$. The $M \times 2$ matrix $\Omega_l$ has entries $(\Omega_l)_{m1} = \int_0^1 \phi_m(t)\, dt$ and $(\Omega_l)_{m2} = \int_0^1 t\, \phi_m(t)\, dt$. The $M \times M'$ matrix $\Omega$ has entries $\Omega_{mm'} = \int_0^1 \phi_m(t) \psi_{m'}(t)\, dt$.

Substituting the basis expansions into the functional trait model gives

$$
\begin{aligned}
y_a &= \int_0^1 \beta\,(t) v_a(t)\,dt + e_a \\
&= \int_0^1 \left\{\beta_{l0} + \beta_{l1} t + \sum_{m\prime=1}^{M\prime} \beta_{m\prime}\, \psi_{m\prime}(t)\right\}\left\{\sum_{k=1}^{K}\sum_{m=1}^{M} w_{km}\, \phi_m(t) g_{ak}\right\} dt + e_a \\
&= \boldsymbol{\beta}_l^\top \Omega_l^\top W^\top \mathbf{g}_a + \boldsymbol{\beta}^\top \Omega^\top W^\top \mathbf{g}_a + e_a.
\end{aligned}
$$

A GWAS estimates the association with each SNP separately. For SNP $k'$, the marginal model is $y_a = b_{k\prime} g_{ak\prime} + \tilde{e}_a$. Because the trait and genotypes are standardized, its regression coefficient is

$$
b_{k\prime} = \mathrm{Cov}(y_a, g_{ak\prime}) = \left(\boldsymbol{\beta}_l^\top \Omega_l^\top W^\top + \boldsymbol{\beta}^\top \Omega^\top W^\top\right)\begin{pmatrix} R_{1k\prime} \\ \vdots \\ R_{Kk\prime}\end{pmatrix},
$$

where $R_{kk\prime}$ is the correlation between genotypes at SNPs $k$ and $k'$, the $(k,\, k')$ entry of the linkage-disequilibrium (LD) matrix $R$. Therefore, the vector of marginal coefficients is $b = RW\Omega_l \boldsymbol{\beta}_l + RW\Omega\boldsymbol{\beta}$.

Following TWiST, we model the GWAS estimates as

$$
\hat{b} \sim N\left(RW\Omega_l \boldsymbol{\beta}_l + RW\Omega\boldsymbol{\beta},\, \frac{R}{N_{\mathrm{GWAS}}}\right), \tag{S3}
$$

where $N_{\mathrm{GWAS}}$ is the GWAS sample size. This model assumes that the GReX of the gene explains a small fraction of the trait variance, as in TWiST.

#### S3. Likelihood based on decorrelated summary statistics

When $R$ is singular because of LD among cis-SNPs, the likelihood in $(\mathrm{S3})$ is a degenerate multivariate normal distribution. Following TWiST, we use the eigendecomposition $R = Q\Lambda Q^\top$, where the columns of the $K \times K'$ matrix $Q$ are orthonormal eigenvectors and $\Lambda$ is the diagonal matrix of the $K'$ nonzero eigenvalues. Applying $Q^\top$ to the summary statistics gives

$$
Q^\top \hat{b} \mid \boldsymbol{\beta} \sim N(\Lambda Q^\top W\Omega_l \boldsymbol{\beta}_l + \Lambda Q^\top W\Omega\boldsymbol{\beta},\, \Lambda/N_{\mathrm{GWAS}}).
$$

As in the main Methods, we use $\boldsymbol{\beta} \sim N(0, \sigma^2 \Omega_2^{-1})$ to control the smoothness of $\beta(t)$, where $\Omega_2$ has entries $\Omega_{2,m\prime n\prime} = \int_0^1 \psi_{m\prime}{}''(t) \psi_{n\prime}{}''(t)\,dt$. Integrating over $\boldsymbol{\beta}$ yields

$$
\begin{aligned}
Q^\top \hat{b} \sim N(\quad & \Lambda Q^\top W \Omega_l \boldsymbol{\beta}_l, \\
& \Lambda/N_{\mathrm{GWAS}} + \sigma^2 \Lambda Q^\top W\Omega\Omega_2^{-1}\Omega^\top W^\top Q\Lambda).
\end{aligned} \tag{S4}
$$

#### S4. Parameter estimation and confidence bands

We follow the TWiST procedure for parameter estimation and confidence bands. To simplify notation, define $D_l = \Lambda Q^\top W \Omega_l$ and $D = \Lambda Q^\top W\Omega$. The variance of the transformed summary statistics is $V = \Lambda/N_{\mathrm{GWAS}} + \sigma^2 D\Omega_2^{-1} D^\top$, and their covariance with the random coefficients is $C = \mathrm{Cov}\big(\boldsymbol{\beta}, Q^\top \hat{b}\big) = \sigma^2 \Omega_2^{-1} D^\top$. For a fixed $\sigma^2$, the maximum likelihood estimate of $\boldsymbol{\beta}_l$ is

$$
\hat{\boldsymbol{\beta}}_l(\sigma^2) = \left(D_l^\top V^{-1} D_l\right)^{-1} D_l^\top V^{-1} Q^\top \hat{b}.
$$

We substitute this estimate into $(S4)$ and maximize the likelihood over $\sigma^2$. The resulting estimate $\hat{\sigma}^2$ gives $\widehat{\boldsymbol{\beta}}_l = \widehat{\boldsymbol{\beta}}_l(\hat{\sigma}^2)$. For fixed $\sigma^2$, the corresponding variance-covariance matrix is

$$\mathrm{Var}\left(\widehat{\boldsymbol{\beta}}_l\right) = \left(D_l^\top V^{-1} D_l\right)^{-1}.$$

The best linear unbiased predictor (BLUP) of the random coefficients is

$$\widehat{\boldsymbol{\beta}} = CV^{-1}\left(Q^\top \hat{b} - D_l \widehat{\boldsymbol{\beta}}_l\right).$$

Its variance is

$$\mathrm{Var}\left(\widehat{\boldsymbol{\beta}}\right) = CV^{-1}AVA^\top V^{-1}C^\top,$$

where $A = I_{K'} - D_l\left(D_l^\top V^{-1} D_l\right)^{-1} D_l^\top V^{-1}$ and $I_{K'}$ is the $K' \times K'$ identity matrix.

For a given pseudotime $t$, the estimated effect curve is

$$\hat{\beta}(t) = (1, t)\widehat{\boldsymbol{\beta}}_l + \boldsymbol{\psi}(t)^\top \widehat{\boldsymbol{\beta}},$$

where $\boldsymbol{\psi}(t) = \left(\psi_1(t), \ldots, \psi_{M'}(t)\right)^\top$. The two coefficient estimates are uncorrelated for fixed $\sigma^2$, giving

$$\begin{aligned}\mathrm{Var}\{\hat{\beta}(t)\} = \quad & (1, t)\mathrm{Var}\left(\widehat{\boldsymbol{\beta}}_l\right)(1, t)^\top \\ & + \boldsymbol{\psi}(t)^\top \mathrm{Var}\left(\widehat{\boldsymbol{\beta}}\right)\boldsymbol{\psi}(t).\end{aligned}$$

We evaluate these variance expressions at $\hat{\sigma}^2$.

The global, dynamic, and nonlinear tests compare the likelihood in $(S4)$ with the likelihood under the corresponding null hypothesis: $\beta(t) = 0$ for the global test, $\beta(t) = \beta_{l0}$ for the dynamic test, and $\beta(t) = \beta_{l0} + \beta_{l1}t$ for the nonlinear test, which correspond to $\beta_{l0} = \beta_{l1} = \sigma^2 = 0$, $\beta_{l1} = \sigma^2 = 0$, and $\sigma^2 = 0$, respectively. The variance component is on the boundary of the parameter space under these null hypotheses. Following TWiST, we use the mixture reference distributions $0.95\chi_2^2 + 0.05\chi_3^2$ for the global test, $0.95\chi_1^2 + 0.05\chi_2^2$ for the dynamic test, and $0.95\delta_0 + 0.05\chi_1^2$ for the nonlinear test, where $\delta_0$ is a point mass at zero[2,3].

Under their respective asymptotic null distributions, $0.95\chi_2^2 + 0.05\chi_3^2$ and $0.95\chi_1^2 + 0.05\chi_2^2$, the global and dynamic test statistics are mixtures of two continuous distributions, so the usual probability-integral-transform argument gives P-values that are asymptotically Uniform(0,1) under the null and span the full interval $(0,1)$. The nonlinear test statistic's null distribution, $0.95\delta_0 + 0.05\chi_1^2$, instead places a point mass at zero: the correct P-value is $p^{\text{nonlinear}} = 1$ if $\mathrm{LRT} = 0$ and $p^{\text{nonlinear}} = 0.05 \cdot P(\chi_1^2 \geq \mathrm{LRT})$ if $\mathrm{LRT} > 0$, where LRT denotes the likelihood-ratio statistic. Because $\sigma^2$ is estimated here by maximizing over a fixed grid of candidate values that never includes exactly $0$, the fitted statistic is always strictly positive in practice, so only the second branch of $p^{\text{nonlinear}}$ is ever realized; the resulting P-values are compressed into $(0, 0.05]$ rather than spanning $(0,1)$, and so do not satisfy the requirement $P(p \leq u) \leq u$ for $u \geq 0.05$ that a valid input to the Cauchy combination test[4] must satisfy. We therefore exclude the nonlinear test from the Cauchy combination.
